# Estimating epidemiological delay distributions for infectious diseases

**DOI:** 10.1101/2024.01.12.24301247

**Authors:** Sang Woo Park, Andrei R. Akhmetzhanov, Kelly Charniga, Anne Cori, Nicholas G. Davies, Jonathan Dushoff, Sebastian Funk, Katie Gostic, Bryan Grenfell, Natalie M. Linton, Marc Lipsitch, Adrian Lison, Christopher E. Overton, Thomas Ward, Sam Abbott

## Abstract

Understanding and accurately estimating epidemiological delay distributions is important for public health policy. These estimates directly influence epidemic situational awareness, control strategies, and resource allocation. In this study, we explore challenges in estimating these distributions, including truncation, interval censoring, and dynamical biases. Despite their importance, these issues are frequently overlooked in the current literature, often resulting in biased conclusions. This study aims to shed light on these challenges, providing valuable insights for epidemiologists and infectious disease modellers.

Our work motivates comprehensive approaches for accounting for these issues based on the underlying theoretical concepts. We also discuss simpler methods that are widely used, which do not fully account for known biases. We evaluate the statistical performance of these methods using simulated exponential growth and epidemic scenarios informed by data from the 2014-2016 Sierra Leone Ebola virus disease epidemic.

Our findings highlight that using simpler methods can lead to biased estimates of vital epidemiological parameters. An approximate-latent-variable method emerges as the best overall performer, while an efficient, widely implemented interval-reduced-censoring-and-truncation method was only slightly worse. Other methods, such as a joint-primary-incidence-and-delay method and a dynamic-correction method, demonstrated good performance under certain conditions, although they have inherent limitations and may not be the best choice for more complex problems.

Despite presenting a range of methods that performed well in the contexts we evaluated, residual biases persisted, predominantly due to the simplifying assumption that the distribution of event time within the censoring interval follows a uniform distribution; instead, this distribution should depend on epidemic dynamics. However, in realistic scenarios with daily censoring, these biases appeared minimal. This study underscores the need for caution when estimating epidemiological delay distributions in real-time, provides an overview of the theory that practitioners need to keep in mind when doing so with useful tools to avoid common methodological errors, and points towards areas for future research.

**Summary:** *What was known prior to this paper:*

- **Importance of accurate estimates:** Estimating epidemiological delay distributions accurately is critical for model development, epidemic forecasts, and analytic decision support.
- **Right truncation:** Right truncation describes the incomplete observation of delays, for which the primary event already occurred but the secondary event has not been observed (e.g. infections that have not yet become symptomatic and therefore not been observed). Failing to account for the right truncation can lead to underestimation of the mean delay during real-time data analysis.
- **Interval censoring:** Interval censoring arises when epidemiological events occurring in continuous time are binned into time intervals (e.g., days or weeks). Double censoring of both primary and secondary events needs to be considered when estimating delay distributions from epidemiological data. Accounting for censoring in only one event can lead to additional biases.
- **Dynamical bias:** Dynamical biases describe the effects of an epidemic’s current growth or decay rate on the observed delay distributions. Consider an analogy from demography: a growing population will contain an excess of young people, while a shrinking population will contain an excess of older people, compared to what would be expected from mortality profiles alone. Dynamical biases have been identified as significant issues in real-time epidemiological studies.
- **Existing methods:** Methods and software to adjust for censoring, truncation, and dynamic biases exist. However, many of these methods have not been systematically compared, validated, or tested outside the context in which they were originally developed. Furthermore, some of these methods do not adjust for the full range of biases.

*What this paper adds:*

- **Theory overview:** An overview of the theory required to estimate distributions is provided, helping practitioners understand the underlying principles of the methods and the connections between right truncation, dynamical bias, and interval censoring.
- **Review of methods:** This paper presents a review of methods accounting for truncation, interval censoring, and dynamical biases in estimating epidemiological delay distributions in the context of the underlying theory.
- **Evaluation of methods:** Methods were evaluated using simulations as well as data from the 2014-2016 Sierra Leone Ebola virus disease epidemic.
- **Cautionary guidance:** This work underscores the need for caution when estimating epidemiological delay distributions, provides clear signposting for which methods to use when, and points out areas for future research.
- **Practical guidance:** Guidance is also provided for those making use of delay distributions in routine practice.

*Key findings:*

- **Impact of neglecting biases:** Neglecting truncation and censoring biases can lead to flawed estimates of important epidemiological parameters, especially in real-time epidemic settings.
- **Equivalence of dynamical bias and right truncation:** In the context of a growing epidemic, right truncation has an essentially equivalent effect as dynamical bias. Typically, we recommend correcting for one or the other, but not both.
- **Bias in common censoring adjustment:** Taking the common approach to censoring adjustment of naively discretising observed delay into daily intervals and fitting continuous-time distributions can result in biased estimates.
- **Performance of methods:** We identified an approximate-latent-variable method as the best overall performer, while an interval-reduced-censoring-andtruncation method was resource-efficient, widely implemented, and performed only slightly worse.
- **Inherent limitations of some methods:** Other methods, such as jointly estimating primary incidence and the forward delay, and dynamic bias correction, demonstrated good performance under certain conditions, but they also had inherent limitations depending on the setting.
- **Persistence of residual biases:** Residual biases persisted across all methods we investigated, largely due to the simplifying assumption that the distribution of event time within the primary censoring interval follows a uniform distribution rather than one influenced by the growth rate. These are minimal if the censoring interval is small compared to other relevant time scales, as is the case for daily censoring with most human diseases.

*Key limitations:*

- **Differences between right censoring and truncation:** We primarily focus on right truncation, which is most relevant when the secondary events are easier to observe than primary events (e.g., symptom onset vs. infection)—in this case, we can’t observe the delay until the secondary event has occurred. In other cases, we can directly observe the primary event and wait for the secondary event to occur (e.g., eventual recovery or death of a hospitalized individual)—in this case, it would be more appropriate to use right censoring to model the unresolved delays. For simplicity, we did not cover the right censoring in this paper.
- **Daily censoring process:** Our work considered only a daily interval censoring process for primary and secondary events. To mitigate this, we investigated scenarios with short delays and high growth rates, mimicking longer censoring intervals with extended delays and slower growth rates.
- **Deviation from uniform distribution assumption:** We show that the empirical distribution of event times within the primary censoring interval deviated from the common assumption of a uniform distribution due to epidemic dynamics. This discrepancy introduced a small absolute bias based on the length of the primary censoring window to all methods and was a particular issue when delay distributions were short relative to the censoring window’s length. In practice, other biological factors, such as circadian rhythms, are likely to have a stronger effect than the growth rate at a daily resolution. Nonetheless, our work lays out a theoretical ground for linking epidemic dynamics to a censoring process. Further work is needed to develop robust methods for wider censoring intervals.
- **Temporal changes in delay distributions:** The Ebola case study showcased considerable variation in reporting delays across the epidemic timeline, far greater than any bias due to censoring or truncation. Further work is needed to extend our methods to address such issues.
- **Lack of other bias consideration:** The idealized simulated scenarios we used did not account for observation error for either primary or secondary events, possibly favouring methods that do not account for real-world sources of biases.
- **Limited distributions and methods considered:** We only considered lognormal distributions in this study, though our findings are generalizable to other distributions. Mixture distributions and non-parametric or hazard-based methods were not included in our assessment.
- **Exclusion of fitting discrete-time distributions:** We focused on fitting continuous-time distributions throughout the paper. However, fitting discretetime distributions can be a viable option in practice, especially at a daily resolution. More work is needed to compare inferences based on discrete-time distributions vs continuous-time distributions with daily censoring.
- **Exclusion of transmission interval distributions:** Our work primarily focused on inferring distributions of non-transmission intervals, leaving out potential complications related to dependent events. Additional considerations such as shared source cases, identifying intermediate hosts, and the possibility of multiple source cases for a single infectee were not factored into our analysis.

## 1 Introduction

Characterizing the distribution of time between two epidemiological events is essential to understanding the course of an infectious disease epidemic and making clinical and public health decisions. For example, some intervals, such as the incubation period (i.e., the time between infection and symptom onset), provide useful means of summarizing aspects of the course of infection of each infected person (Lauer et al., 2020; Linton et al., 2020; Verity et al., 2020a). Other intervals, such as the generation interval (i.e., the time between infection and transmission) and serial interval (i.e., the time between symptom onsets in a transmission pair), describe the transmission pattern across multiple individuals (Madewell et al., 2023) and can provide information about how infectiousness varies over the course of an infection (Sender et al., 2022). Combining these intervals can further help determine the controllability of epidemics (e.g., potential for pre-symptomatic or asymptomatic transmission (Fraser et al., 2004)) and inform guidelines for intervention measures, such as isolation of cases (Hellewell et al., 2020), and travel screening (Gostic et al., 2020). Other intervals, such as reporting delays (e.g., the time from symptom onset until a case is reported), are needed for real-time data interpretation, analysis, and for epidemic forecasting (Marinović et al., 2015; Overton et al., 2022; Abbott et al., 2020; Beesley et al., 2022). Biases in delay distribution estimates can therefore translate to biases in the chain of evidence used to inform decisions (Lipsitch et al., 2020).

There are multiple sources of bias that can affect the estimation of epidemiological delay distributions. Some of these pertain to data reliability issues, such as recall bias, whereas others are intrinsically linked to the structure of the data collection process. In this paper, we primarily focus on the latter type of bias.

First, event observations are very often *censored*, meaning that we don’t know when the event happened exactly but we do know it occurred. Instead, epidemiological events are typically reported using an interval (e.g. a date, a week, or a range of dates) rather than the exact time at which an individual experienced an event (Lindsey and Ryan, 1998). This is known as *interval censoring* as we only know the interval in which the event occurred. Only on rare occasions, the time of the event known is known more precisely (i.e. to the hour, minute, or very rarely to the second), such as the time of death recorded on a death certificate, but even in these cases uncertainty often remains.

Interval censoring can be particularly problematic if the reporting interval is relatively wide compared to the typical length of a delay. This is a common problem for short delays: for example, influenza has short generation intervals (2–3 days) and incubation periods (1–2 days) (Fraser et al., 2009), meaning that even daily censoring is expected to be problematic. Even when the delays are extremely long (e.g., the incubation periods for TB or HIV/AIDS), interval censoring can be problematic because the widths of censoring intervals are also just as wide.

Even when interval censoring is taken into account, many studies only adjust for the censoring of a single event, rather than both primary and secondary events. Examples of this include accounting for censoring of the date of exposure but not the date of symptom onset when estimating the incubation period (Backer et al., 2020, 2022). In addition to interval censoring, data can be either left-or right-censored, where only the upper or lower bounds, respectively, of the event times are known. In the case of left censoring, we can always put a realistic lower bound, such as the beginning of an epidemic, meaning that standard methods for interval censoring can be readily applied. On the other hand, right censoring typically corresponds to the case where we have already observed the primary event but failed to observe the secondary event, typically due to a dropout of a patient from a study and failure to follow up. Multiple methods have been developed for modelling right-censoring (Ghani et al., 2005).

Second, epidemic data often suffer from *right truncation*, meaning that we only observe events that have already happened and been reported (Brookmeyer and Damiano, 1989; Kalbfleisch and Lawless, 1989; Gelman et al., 2013). In contrast to right censoring, for which we have a partial observation of the lower bound of the secondary event, we do not have any information in the case of truncation. For example, incubation periods are often truncated because we are unable to observe the primary event (infection) directly until the secondary event (symptom onset) is reported. Failure to correct for right truncation can bias the data toward observation of shorter intervals (e.g. if only individuals with symptoms that resolved before the end of a study are included in the analysis). Moreover, only a limited number of studies consider the interaction between censoring and truncation (Linton et al., 2020; Ward and Johnsen, 2021).

Finally, recent studies have highlighted the role of dynamical biases: during the growth phase of an epidemic, we are more likely to observe shorter delays because a disproportionately large number of individuals have been infected more recently; this effect is reversed during epidemic decay (Britton and Scalia Tomba, 2019; Park et al., 2022). The effects of dynamical biases on the observed delay distributions are quantitatively equivalent to truncation biases during the exponential growth phase, but their equivalence has not always been clear, leading to attempts to address both biases simultaneously (Linton et al., 2020; Guo et al., 2023b; Verity et al., 2020c). These approaches further highlight that clearer guidance, and more robust methods that take account of this guidance, are needed to handle biases found in different epidemiological contexts, which can depend on the data-collection method, type of data (e.g., single-individual delays such as incubation periods vs pair-dependent delays such as generation or serial interval), and underlying epidemic dynamics.

Several methods and software packages accounting for censoring and truncation corrections in epidemiological data already exist. The recent COVID-19 and mpox epidemics have seen a marked increase in the development of both methods and software implementations (Backer et al., 2020; Linton et al., 2020; Tindale et al., 2020; Verity et al., 2020a; Hart et al., 2021). However, many of these methods were developed for a particular context, have not been validated sufficiently, or do not cover the full range of potential biases. Developed in 2009, `CoarseDataTools` is widely used (Thompson et al., 2019; Madewell et al., 2023). It provides methods that can account for double censoring (censoring of both primary and secondary events) (Reich et al., 2009, 2010). However, it does not account for truncation bias, and was not implemented to be readily extensible. Part of its popularity is driven by its implementation in the popular effective-reproduction-number-estimation package `EpiEstim` for serial-interval estimation (Cori et al., 2013; Thompson et al., 2019). Unfortunately, naively relying on this implementation may lead to biased effective reproduction number estimates.

Other widely used examples include the method of Backer et al. (2020), which allows for uniform censoring in the primary event using latent variables but does not account for right truncation. Similarly, the method of Linton et al. (2020) allows for double censoring as well as right truncation adjustment. The method of Ward et al. (2022) also accounts for double censoring and right truncation. However, so far, these approaches have not been validated against simulations.

Tools exist for analyzing non-domain-specific interval-censored data (see Pan et al. (2020) for a detailed review and comparison), but only a few of them account for right truncation and these methods are rarely used within the infectious disease modelling community. The lack of ready-to-use software implementations, or standardised methods, that can adjust for both censoring and right truncation means that many of the estimated delay distributions present in the literature are likely biased. An additional issue is the predominance of methods developed specifically for the application in which they were used (Backer et al., 2020; Linton et al., 2020; Guo et al., 2023b), which means that even when known biases appear to have been accounted for this can often be hard to verify due to the lack of robust evaluation. Estimates from early in epidemics are likely particularly biased due to unaccounted-for right-truncation, while retrospective estimates, though likely less biased, may still be problematic when censoring is not properly taken care of.

In this work, we aim to provide clear methodological and practical guidance for researchers tasked with estimating epidemiological delay distributions. We also aim to provide robust and flexible tools and methods, both rederivations of those presented elsewhere and novel, for them to apply this guidance in practice. We do this by first introducing some general theories for characterizing epidemiological delay distributions justified by grouping individuals into cohorts based on their observation time. We then introduce in detail the biases that are common when estimating epidemiological delay distributions. Based on this understanding, we then introduce an exact method for accounting for common biases. We also introduce a range of other commonly used and novel methods that represent simplifications of this approach. We then evaluate these methods, both on simulated scenarios and on data from the 2014-2016 Sierra Leone Ebola Virus disease epidemic. Finally, we consider areas that require further development. Whilst the details of each section are important, we also provide a summary for each that contains the main points in order to aid understanding. In addition, we have summarised what was known prior to this paper, what this paper adds, key findings, and key limitations.

## 2 Methods

### 2.1 General theory for measuring epidemiological delay distributions

Here we give a conceptual, visual, and mathematical overview of the general theory for measuring epidemiological delay distributions and the specific theory relating to dynamic bias. This theory is then used in later sections to relate to other forms of bias and to justify and develop inference methods. We first give a summary of the key points.

#### Summary

- The *intrinsic* distribution is the theoretical distribution that characterises the underlying epidemiological process of interest. It describes the probability of waiting a certain amount of time between a *primary* and a *secondary* event (e.g. between infection and symptom onset) under constant conditions in the population.
- Realised epidemiological delays can be measured both forward, starting from the primary event toward the secondary event, or backward, starting from the secondary event back to the primary event. For any given individual, the direction of measurement does not affect the length of the delay. Intrinsic and realized transmission intervals (e.g., generation interval and serial interval) can differ systematically due to changes in transmission conditions (e.g., susceptible pool in the population).
- The *forward distribution* is measured from a cohort of individuals who experienced the primary event at the same time and is expected to give a good estimate of the intrinsic distribution when conditions remain constant. For modelling purposes, the forward distribution is often preferred over the backward distribution as it better approximates the intrinsic distribution.
- The *backward distribution* is measured from a cohort of individuals who experienced the secondary event at the same time. For a given intrinsic distribution, the backward distribution can systematically vary over time and differ from the forward distribution. This is due to the interaction between the observation process and the temporal change in the incidence of the primary event.

##### Conceptual overview

For any epidemiological process, there is an underlying *intrinsic* distribution that describes the time difference between events in the process. In the context of epidemiological delays, the intrinsic distribution is a theoretical distribution that describes the probability of waiting a certain amount of time between a *primary* and a *secondary* event (e.g. between infection and symptom onset) under constant conditions in the population (Champredon and Dushoff, 2015). This distribution can be calculated by averaging across individuals: for example, the intrinsic generation-interval distribution only depends on the average infectiousness of infected individuals at a given time and does not depend on other factors, such as intervention measures or the proportion susceptible (Park et al., 2021). As the intrinsic distribution characterises the underlying infection characteristic, it is used for modeling, and is therefore often the distribution that we want to estimate. However, the intrinsic distribution is generally not directly observable, as it may differ from the realised distributions that are measured from the actual primary and secondary events observed during an epidemic. In particular, realised distributions can change over the course of an epidemic: for example, changes in realised generation intervals can reflect changes in transmission dynamics, including susceptible depletion. For practitioners, characterizing both the intrinsic and realised distribution is important. In most cases, the primary event always occurs before the secondary event (e.g., infection followed by symptom onset). However, there are exceptions: for example, an infectee may develop symptoms before their infector (Svensson, 2007). Negative delays can also happen for single-individual events: an infected individual may test positive before or after symptom onset (Singanayagam et al., 2020). Here we focus on non-negative, single-individual delays, i.e. where the secondary event always happens after the primary event. We also focus on the case where forward distributions do not change over the course of an epidemic—later in the Ebola virus disease example, we show that this is not always the case. Many of the lessons in this paper will however carry over to more complicated cases involving negative delays and/or delays between epidemiological events from two individuals.

We take the reporting delay—defined as the time from symptom onset (the primary event) to confirmation (the secondary event)—as an example. There are two different approaches to measuring this interval. If we group individuals based on their time of symptom onset and follow them until confirmation we are measuring *forward* delay. If we instead group individuals based on when they tested positive and ask when they developed symptoms, we are measuring *backward* delays. For any individual realisation of a delay as a paired primary and secondary events, the length of forward and backward delays are identical. However, in a *cohort* of individuals that experienced the primary (forward) or secondary (backward) events at the same time, the resulting forward and backward distributions can systematically differ when incidence is changing over time. Taking the cohort approach also allows us to ask how the forward and backward distributions change over the course of an epidemic.

As an example, consider a population where the incidence of infection is growing exponentially (Fig. 1A). If we take a cohort of individuals who developed symptoms on day 25, then we observe Fig. 1B, which corresponds to a *forward* distribution. In this case, we see that the forward distribution in Fig. 1B matches the intrinsic distribution that we used to simulate Fig. 1A. In general, we expect the forward distribution to approximate the intrinsic distribution reasonably well, in particular under relatively stable external conditions (Park et al., 2021). This means that the forward distribution (rather than the backward distribution) is most useful for downstream analysis, modeling, and decision support.

**Figure 1:**
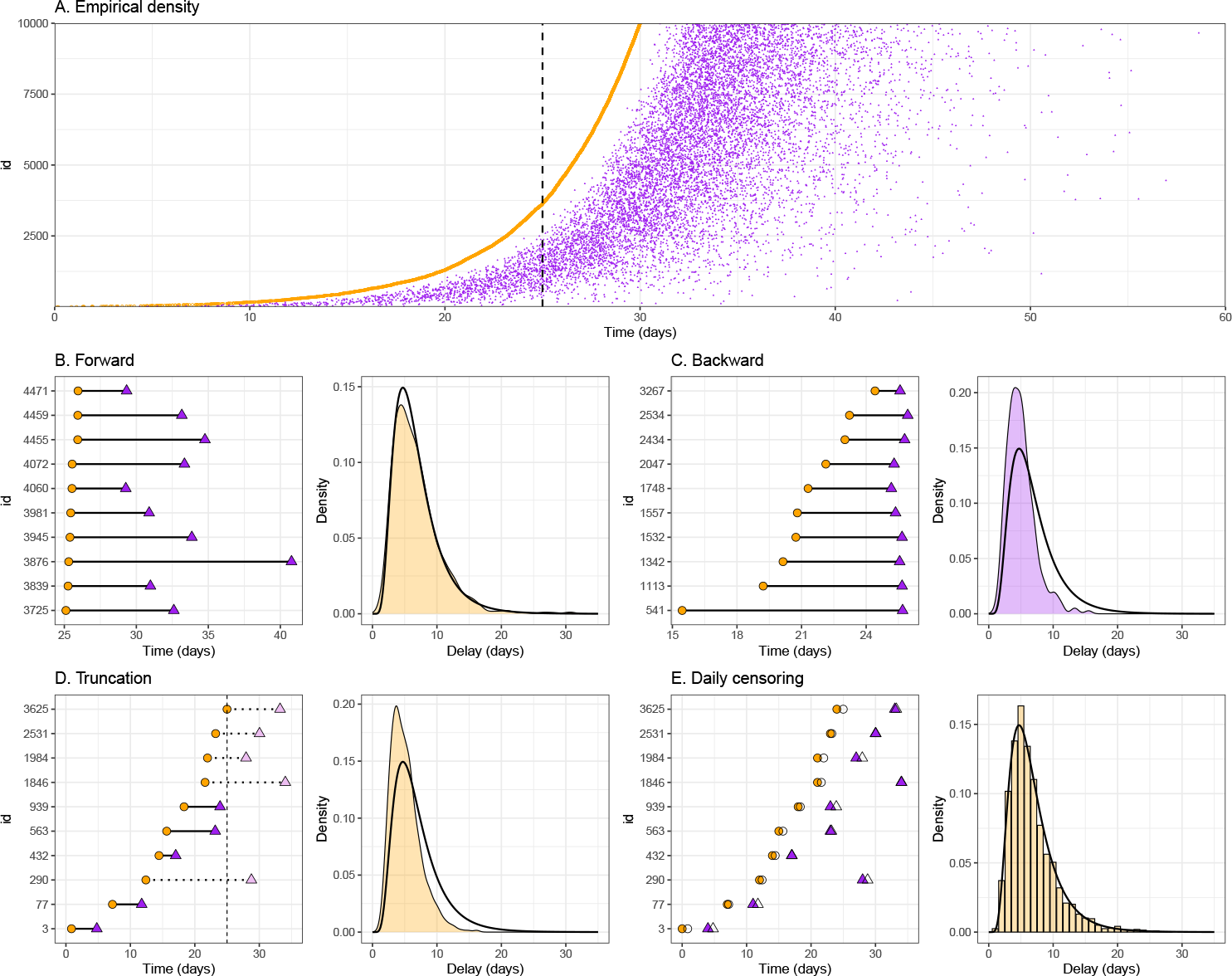
Schematic diagrams showing forward and backward distributions with the impact of truncation and censoring biases. (A) The density plot of exponentially increasing cumulative cases of primary events (orange) and the corresponding secondary events (purple). Case IDs (y-axis) are ordered by the primary event time. The dashed vertical line indicates day 25. Panels B–E show four different delay distributions using day 25 as a reference day. The left panels show a sample data set with circles representing the timing of primary events and triangles representing the timing of secondary events. The right panels show the corresponding distributions (filled areas) against the intrinsic distribution (transparent areas, solid lines). (B) The empirical forward distribution measured from a cohort of individuals who experienced the primary event on day 25. (C) The empirical backward distribution measured from a cohort of individuals who experienced the secondary event on day 25. (D) The truncated distribution measured from all individuals who experienced both the primary and secondary events before day 25 (i.e., those with solid lines connecting their events). (E) The observed distribution under daily reporting from all individuals who experienced the primary event before day 25 (without truncation). Empty circles and triangles represent the event dates under daily reporting.

Some delay distributions are observed when the secondary event is reported, and the timing of the primary event is then recalled or inferred (e.g. in a reporting delay, typically the confirmation is first observed and the date of symptom onset recalled by the patient). Since at this point, both primary and secondary events have already happened, backward-looking cohorts, and hence backward distributions, typically do not have truncation. For example, if we take a cohort of individuals who tested positive on day 25, we observe Fig. 1C which corresponds to a *backward* distribution. In this case, we see that the backward distribution has a shorter mean than both the intrinsic and forward distributions. This is because the backward-looking cohort contains a mixture of individuals who developed symptoms recently (shorter reporting delays) and individuals who developed symptoms further in the past (longer reporting delays). In the case of a growing epidemic, individuals who developed symptoms recently are more abundant, and so the backward distribution is shifted toward shorter intervals. Similarly, when the epidemic is declining, individuals who developed symptoms recently are rare, and so the backward distribution is shifted toward longer intervals (and has a longer mean than the intrinsic distribution) (Xin et al., 2021). More generally, the backward distribution always depends on the timevarying incidence of primary events. We refer to this dependence as dynamical bias because it biases the backward distribution in comparison to the forward distribution (Park et al., 2021).

Additionally, the realized forward and backward distributions are both susceptible to a range of biases. Both can depend on events that are censored and both may be truncated. In the case of forward-looking cohorts, the most common bias is right truncation as the data set may be finalized for analysis before all individuals are observed. Similarly, the backward distribution suffers from left truncation when data on primary events are not available before some time point (Cain et al., 2011).

In particular, we note that, in the exponential growth case, there is a theoretical equivalence between backward and truncated forward cohorts. This can be seen in Fig. 1D, which can be calculated by taking all individuals who were infected and developed symptoms before day 25 (therefore truncating all events that happened after day 25). This results from an equivalence of the backward and truncated cohorts in exponential growth settings. In the following section, we describe the effects of truncation (including its similarities and differences with the dynamical bias) and censoring in detail.

##### Mathematical definition

To sharpen our discussion of forward and backward distributions, we define them mathematically. As in the conceptual overview and Fig. 1, we often define cohorts based on time intervals (e.g., a group of individuals who were infected on the same day, same week, or since the start of an epidemic). Mathematically, we can instead define incidence, and therefore cohorts, at any given, infinitely precise, *time point* : for example, in the standard continuous-time Susceptible-Infected-Recovered (SIR) model, the incidence of infection at time *t* corresponds to *βS*(*t*)*I*(*t*), where *β* represents the transmission rate, *S* represents the number of susceptible individuals, and *I* represents the number of infected individuals.

Here, we write *f*_*p*_(*τ*) to represent the conditional probability density of observing a forward delay of length *τ* given that the primary event occurred at time *p*. Whilst the forward distribution can vary, for example over time or by risk or age groups, we primarily focus on cases where the forward distribution remains stable over the course of an epidemic (i.e., *f*_*p*_(*τ*) = *f* (*τ*) for all *p* in this paper), and is thus identical to the intrinsic distribution. We also write 𝒫(*p*) to denote the incidence of primary events (i.e., the rate at which individuals experience the primary event at time *p*). Likewise, we write *b*_*s*_(*τ*) to represent the conditional probability density of observing a backward delay of length *τ* given that the secondary event occurred at time *s*. We also write 𝒮(*s*) to denote the incidence of secondary events (i.e., the rate at which individuals experience the secondary event at time *s*). Then, the total density of individuals 𝒯(*p, s*) who experienced the primary event at time *p* and secondary event at time *s* can be equivalently expressed in terms of the forward distribution:

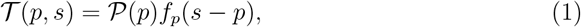

and the backward distribution:

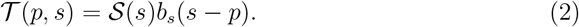

We can then write down a relationship between the forward and backward distributions using the above relationship:

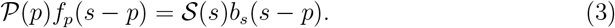

By substituting *p* = *s*− *τ* and rearranging, we see that the backward distribution is given by the incidence of primary events normalized by the size of the relevant cohort:

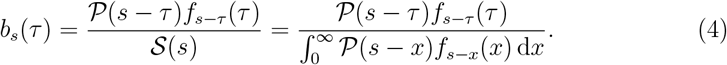

Here, the denominator 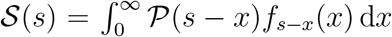 represents the normalization constant such that the probability distribution integrates to 1. In particular, when the incidence of primary events is growing exponentially at rate *r* and the forward distribution is static (i.e. *f*_*p*_(*τ*) = *f* (*τ*)), the backward distribution is also static, and given by:

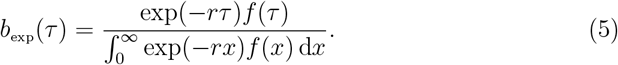

This simplified relationship has been used extensively in the literature (Britton and Scalia Tomba, 2019; Verity et al., 2020b; Park et al., 2022). When *r >* 0, the backward distribution will have a shorter mean than the corresponding forward distribution because we are more likely to observe individuals who experienced the primary event recently. This dependence on the incidence of the primary event leads to *dynamical bias* when the backward distribution is used as an estimate of the intrinsic distribution (Park et al., 2022). As can be seen from Eq. (5), the forward and backward distributions will be equivalent when incidence is stable (*r* = 0).

### 2.2 Biases in estimating forward distributions

In this section, we lay out mathematical foundations for understanding truncation and censoring biases. The framework described in this section will provide a basis for deriving likelihoods for inference in the following section. We first give a summary of the key points.

#### Summary

- Right truncation occurs because we cannot observe event pairs whose secondary event lies in the future. We do not know how much of the data is truncated until all event pairs have been fully observed. This effect biases us toward observing shorter delays when working in real time.
- The amount of right truncation increases as the epidemic grows faster because the amount of recent primary events becomes relatively more common.
- When incidence is growing exponentially, the effect of right truncation on the forward distribution is quantitatively equivalent to the effect of dynamical bias on the backward distribution. When incidence is shrinking, this is not the case: there will be minimal right truncation but the dynamical bias causes the backward distribution to have a longer mean.
- Censoring occurs when the precise time of an event is unknown but we know an event has occurred. Here, we focus on fitting continuous-time distributions. This means that the recording of the exact *date* of an event is implicitly censored because the exact *time* of an event (within a given date) is still unknown. Alternatively, it would be possible to model this using discrete-time distributions—we do not explore this approach in this paper.
- When we estimate delay distributions, we have to account for censoring in both primary and secondary events. Typically, many researchers rely on a single-day censoring window, which can give biased inferences.
- Assumptions about event time distributions within a censoring interval can bias the estimation of continuous-time distributions from the observed discrete-time delays as well as the discretisation of continuous-time distributions. Modeling choices for fitting continuous-vs discrete-time distributions need to be carefully considered.

##### Right truncation

Right truncation arises from the inability to observe secondary events that happen after the observation time *T* (see Fig. 1D). For example, during an ongoing epidemic, we tend to underestimate the infection fatality risk because we cannot observe all of the outcomes for infected individuals in real time due to the delay between infection and death (or recovery) and because we typically base observation on individuals having died (or recovered) (Ghani et al., 2005; Lipsitch et al., 2015). In general, we don’t know how much of the data is truncated until all outcomes have been fully observed (Gelman et al., 2013, p. 227). Since right truncation limits our ability to observe long intervals (i.e. those in the right tail of a distribution), it will generally bias us towards observing shorter delays (and therefore to underestimate the mean).

To define this mathematically, we introduce random variables *P* and *S* representing primary and secondary events. Then, the conditional probability of observing a forward delay of length *τ* (i.e., *S* = *P* + *τ*) given primary event time *P* = *p* and observation time *T* (i.e., *S < T*) can be written as:

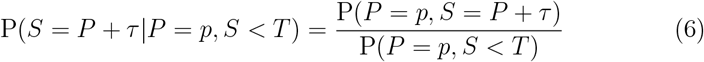

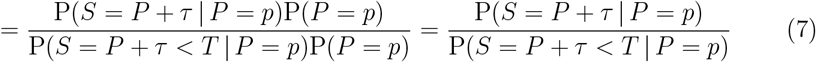

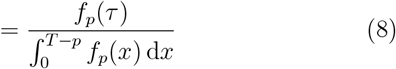

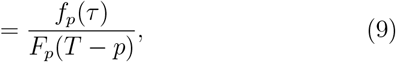

where *f*_*p*_ is the forward distribution and *F*_*p*_ is the forward cumulative probability distribution. Conceptually, Eq. (9) is saying that the probability of observing a given delay from a truncated distribution is equal to the probability of observing that delay from the untruncated forward distribution normalised by the probability of observing any delay before the end of observation. From a cohort perspective, when we measure forward delays from individuals who experienced primary events at time *p < T*, we can only observe delays that are shorter than *T*− *p*. This means that the amount of truncation is higher in a cohort of individuals who experienced their primary events more recently.

The population-level effect of right truncation will be exaggerated when an epidemic is growing because there will be more individuals who experienced primary events recently, meaning their secondary events may not have happened yet. When growth is exponential, the effect of right truncation is quantitatively equivalent to dynamical bias, as shown in Fig. 1C–D. In contrast, when the epidemic is declining quickly, fewer individuals experienced primary events recently, and the effect of right truncation can become negligible as few delays are unobserved. On the other hand, dynamical bias will still be an issue because a cohort of individuals who experienced secondary events during the decay phase will include a relatively large proportion of those who had their primary events further in the past, causing the backward distribution to have a longer mean than the forward distribution. As dynamical biases specifically refer to the dependence of backward distribution on epidemic dynamics, they need not be taken into account when analyzing the forward distribution as long as biases due to truncation are considered.

##### Interval censoring

Censoring occurs when the exact event time is unknown. In contrast to truncation, we know that the focal event happened in the case of interval censoring; we just do not know precisely when. For example, we know that a symptomatic individual was infected sometime before symptom onset but we usually don’t know when. Even if the exact date of the event is known, implicit censoring remains because the event could have theoretically occurred at any time within a 24-hour time window. This implicit censoring discretises the observed delay distribution (Fig. 1E). To define censoring of both primary and secondary events, which is known as double censoring (Reich et al., 2009), we start by first defining the simple single censoring case. Here the secondary event is censored between time *S*_*L*_ and *S*_*R*_, and the primary event (*p*) is known. Then, we have the following relationship,

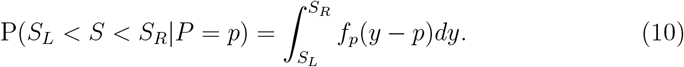

Integrating this leaves

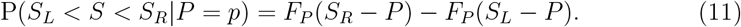

This relationship is commonly used in practice to discretise delay distributions, particularly into daily intervals (Flaxman et al., 2020; Abbott et al., 2020). However, as we have conditioned on the primary event being known, this approach is not appropriate for the common case where *P* is also censored, for example, if only a date is known. The degree of bias this will introduce depends on event probability within the primary censoring interval.

For the more common setting where the primary event is also censored between time *P*_*L*_ and *P*_*R*_ the conditional probability that the secondary event occurs between time *S*_*L*_ and *S*_*R*_ can be written as:

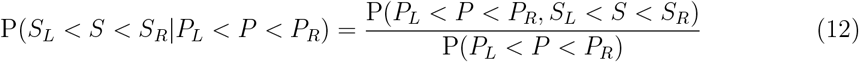

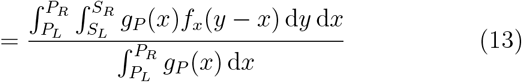

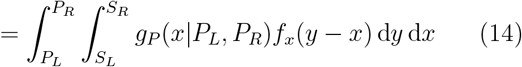

Here, *g*_*P*_ (*x*) is the probability distribution of the primary event time, and *g*_*P*_ (*x*| *P*_*L*_, *P*_*R*_) is the conditional probability distribution of the primary event time given its lower and upper bounds,

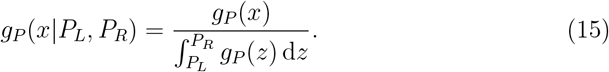

As *g*_*P*_ (*x*) depends on the incidence of primary events, and therefore epidemic dynamics, this can be rewritten as,

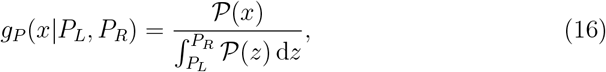

where *P* (*x*) is a continuous-time incidence, as explained earlier; in practice, accounting for this factor will require estimating continuous-time incidence from discrete-time incidence. Making the simplifying assumption of a fixed epidemic growth rate *r*, the conditional probability can then be rewritten as,

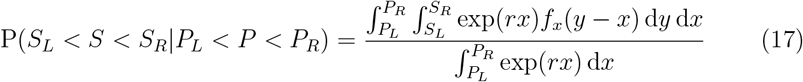

The shape of the conditional probability distribution *g*_*P*_ (*x*|*P*_*L*_, *P*_*R*_) is shown in Fig. 2 for different epidemic growth rates.

**Figure 2:**
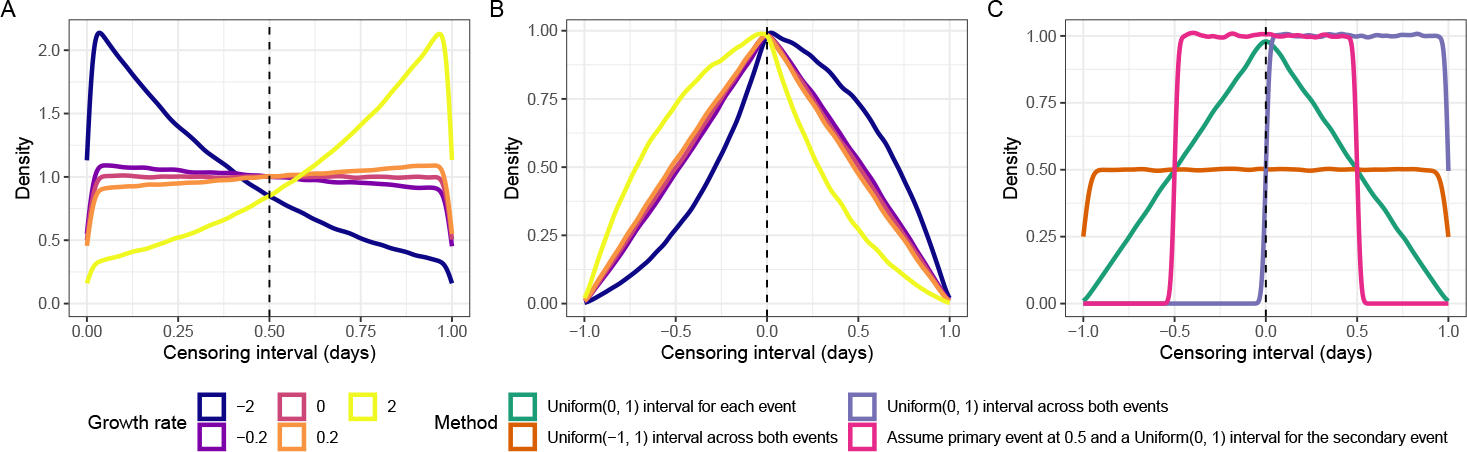
Schematic diagrams showing different assumptions about interval censoring. (A) The distribution of primary event times within a one-day censoring interval across different growth rates. Note that a sudden decrease in probability densities around 0 and 1 are numerical artifacts from plotting. (B) The corresponding distribution of weights for the interval-reduced censoring, which is a convolution between the distribution of event times of primary and secondary events within each censoring interval. (C) The distribution of weights for the interval-reduced censoring for different approximations.

##### Censoring and truncation

Finally, to understand the joint effects of right truncation and interval censoring on the forward distribution, we can quantify the conditional probability of the secondary event occurring between time *S*_*L*_ and *S*_*R*_ given the primary event occurring between *P*_*L*_ and *P*_*R*_ and the observation time *T* :

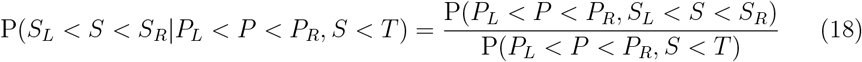

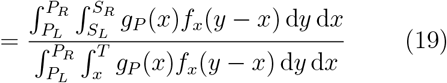

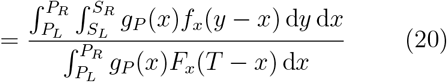

By dividing both the numerator and denominator by 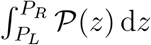, we have:

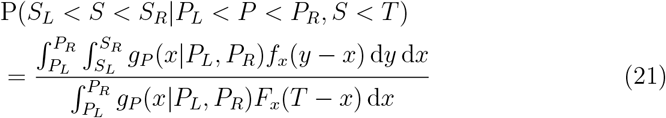

Here, expressing the probability distribution of primary events *g*_*P*_ (*x*) in terms of the conditional probability distribution *g*_*P*_ (*x*|*P*_*L*_, *P*_*R*_) allows for a more flexible inferential framework later on. We also see that the truncation problem also depends on the censoring problem: uncertainties in the primary event time further affect the amount of truncation. This framework generalises the work of Seaman et al. (2022) who reviewed various methods for dealing with right truncation bias but did not account for interval censoring.

### 2.3 Methods for estimating forward delay distributions from observed data

In this section, we describe a method for estimating delay distributions from the observed data based on the theory for censoring and truncation. We then introduce a series of approximations in order to motivate commonly used methods from the literature in order of least to most approximate (with a corresponding decrease in their ability to account for right truncation and censoring biases).

#### Summary

- There is an exact solution for modelling delays that are double censored and right truncated. In practice, this method is not generally practical or stable in real-time settings, and for this reason, we discuss a series of approximations in use.
- Many of the more commonly used approximations do not fully account for censoring, truncation, or both forms of bias. There are also a range of tradeoffs that need consideration.
- Methods that use a latent variable censoring approach and adjust for the observation time are expected to be the most robust currently available for real-time estimation.

##### 2.3.1 Simplifying assumptions for the following methods

We only consider single-individual delays, where we have independent observations of event times across different individuals. This means that in this work, we do not consider methods that account for non-independence in generation or serial intervals. These pair delays also introduce additional biases due to uncertainties about who infected whom—we do not address these issues here. Nonetheless, the methods we present here can still be applied to adjust for truncation, censoring, and dynamical biases in estimating generation-and serial-interval distributions. Finally, we also assume that the forward distribution stays constant over time (i.e., *f*_*p*_(*τ*) = *f*_forward_(*τ*)). All of the methods we consider can be generalised to delays that vary in discrete time using our implementations.

##### 2.3.2 Exact censoring and truncation method

Under daily reporting, the exact timing of the primary and secondary event times are unknown. Instead, the corresponding observed event times are *P*_*L*_ = ⌊*P* ⌋ and *S*_*L*_ = ⌊*S*⌋, respectively. Likewise, the corresponding upper bounds for the censoring interval are *P*_*R*_ = *P*_*L*_ + 1 and *S*_*R*_ = *S*_*L*_ + 1, respectively. Following Eq. (21), the joint likelihood can be written as:

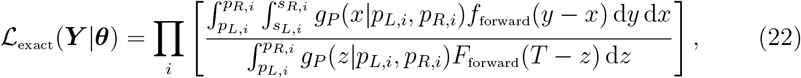

where ***θ*** represents the parameter vector, and ***Y*** represents the data vector.

From a practical standpoint, it is difficult to use this likelihood for inference, especially in real-time epidemic monitoring applications where estimation must be fast and robust. Solving double integrals analytically may be impossible, and calculating them numerically can be computationally costly and numerically unstable. Instead, we can implement this model using an equivalent Bayesian latent variable approach. Here we can treat the primary and secondary event times as latent variables (denoted *x*_*i*_ and *y*_*i*_, respectively), conditional on the known upper and lower bound of these events for each individual, and then integrate across the uncertainty. Then, by taking away both integrals in the numerator, Eq. (22) can be re-written as:

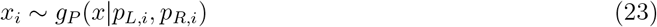

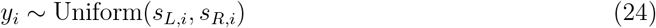

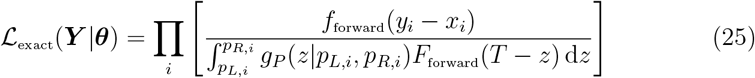

Here, we use the uniform distribution for the secondary event time *y*_*i*_, so that *y*_*i*_ does not affect the likelihood, which allows Eq. (25) to be equivalent to Eq. (22). Using any other distribution for *y*_*i*_ will cause Eq. (25) to deviate from Eq. (22) and therefore will be incorrect.

In practice, modeling the conditional distribution of primary event time *g*_*P*_ (*x*| *p*_*L,i*_, *p*_*R,i*_) is expected to be a difficult problem as it depends on the changes in the incidence of primary events (Eq. (16)). However, if we assume that the incidence of the primary event is changing at a fixed growth rate *r* within the censoring interval we can further simplify the problem:

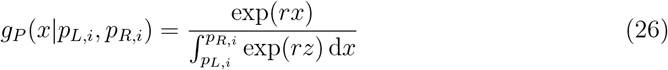

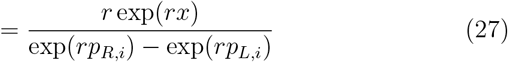

Generalizing the problem beyond stable exponential growth within the interval requires a way of modeling realistic changes in incidence. Even with this simplifying assumption in most real-world settings this would require joint estimation of the growth rate of the primary events in order to properly propagate uncertainty; however, such a model is likely to be computationally more costly.

The exponential scenario provides insights for understanding the method introduced by Linton et al. (2020) during the beginning of the SARS-CoV-2 pandemic. Under Eq. (27), our likelihood is similar in form to the likelihood presented in Linton et al. (2020) with one major distinction (among several other minor differences). The conditional probability *g*_*P*_ (*x p*_*L,i*_, *p*_*R,i*_) we present here converges to a uniform distribution as *r*→ 0 as expected when incidence is stable (Fig. 2A). On the other hand, the corresponding component in Linton et al. (2020) is modeled as *r* exp(−*ru*)*/*(1 − exp(−*ru*)), where *u* is a variable of integration; this term does not converge as *r* → 0.

##### 2.3.3 Approximate latent variable censoring and truncation method

When the primary event time is assumed to be uniformly distributed and censoring interval is narrow, the integral in the denominator of Eq. (25) 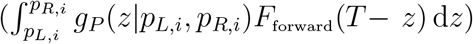 can be approximated by *F*_forward_(*T − x*_*i*_) for some *p*_*L,i*_ *< x*_*i*_ *< p*_*R,i*_. Under these conditions, the exact method can be rewritten as (Ward et al., 2022):

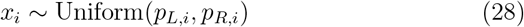

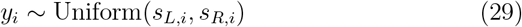

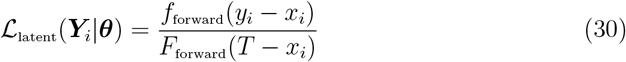

The likelihood for this method corresponds to the conditional probability under right truncation (Eq. (9)). This is a convenient approximation as it does not require additional integration. While it would be possible to extend this method to account for more complex prior distributions that capture epidemic growth or daily activities (e.g., circadian rhythms), the exact method (Eq. (25)) should be considered to avoid potential biases arising from the approximations.

##### 2.3.4 Interval-reduced censoring and truncation methods

Without using latent variables, we can equivalently write down the approximate latent variable model as follows:

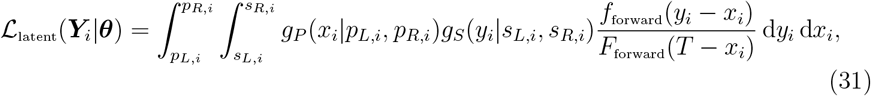

where *g*_*S*_(*y*_*i*_|*s*_*L,i*_, *s*_*R,i*_) corresponds to the uniform distribution between *s*_*L,i*_ and *s*_*R,i*_. When we rewrite in terms of the delay, *d*_*i*_ = *y*_*i*_ − *x*_*i*_, we know that *d*_*i*_ should range between *s*_*L,i*_ − *p*_*R,i*_ and *s*_*R,i*_ − *p*_*L,i*_. Further assuming *F*_forward_(*T* − *x*_*i*_) ≈ *F*_forward_(*T*− *p*_*R,i*_), we can write the above likelihood using a single integral:

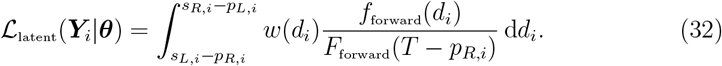

Here, *w*(*d*_*i*_) is a convolution between *g*_*P*_ (*x*_*i*_|*p*_*L,i*_, *p*_*R,i*_) and *g*_*S*_(*y*_*i*_||*s*_*L,i*_, *s*_*R,i*_):

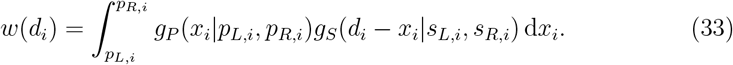

When both *g*_*P*_ (*x*_*i*_|*p*_*L,i*_, *p*_*R,i*_) and *g*_*S*_(*d*_*i*_ − *x*_*i*_|*s*_*L,i*_, *s*_*R,i*_) are assumed to follow uniform distributions, *w*(*d*_*i*_) will have a trapezoid shape (Fig. 2C), as discussed in Reich et al. (2009).

In Fig. 2B–C, we show how the shape of the weight function *w*(*d*_*i*_) changes fordifferent epidemic growth rates (B) and for different assumptions (C). In particular, assuming a uniform distribution for both events censoring intervals approximates the interval-reduced censoring window well when the growth rate is small compared to the width of the censoring window and less well when this is not the case. In Fig. 8, we present the consequences of different assumptions about *w*(*d*_*i*_) on the resulting continuous-time distributions.

We can further simplify the likelihood by assuming a uniform distribution for *w*(*d*_*i*_) (Fig. 2C):

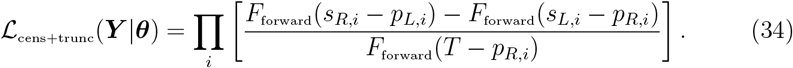

This method is simple to implement as well as being computationally convenient and supported by many software packages, such as the `brms` package (Bürkner, 2018) in the `R` programming language (R Core Team, 2019).

However, it does not fully account for the censoring bias as the uniform assumption across the combined interval is not a good match for the prior that results from combining two uniforms or in more complex settings a growth-mediated prior and a uniform (Fig. 2B–C) or the impact of censoring on the amount of truncation due to the simplified denominator. This means that, depending on the amount of censoring and the shape of the delay distribution, this method will have a residual bias in both the mean and standard deviation of the estimated distribution (Fig. 8). In the special case when the primary event time is exactly known (therefore, *p*_*L,i*_ = *p*_*i*_ = *p*_*R,i*_), this method would be accurate, and also equivalent to the latent variable method.

In the daily reporting setting, we consider in the rest of this paper, the likelihoodcan be reformulated in terms of the observed, discrete-time delay (*d*_i_ = *S*_*L,i*_ − *P*_*L,i*_):

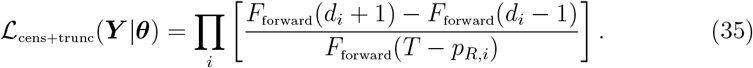

Rephrasing the likelihood in this way highlights that the correct censoring interval for this approach is [*d*_*i*_ − 1, *d*_*i*_ + 1]. Instead, many researchers tend to rely on the [*d*_*i*_, *d*_*i*_ + 1] interval (Flaxman et al., 2020; Abbott et al., 2020), which in effect only accounts for the censoring of a single event and hence induces additional bias beyond that introduced by assuming a uniform distribution. Other potential approximations are [*d*_*i*_ − 0.5, *d*_*i*_ + 0.5], which assumes the primary event is known to have taken place at the midpoint of the interval, and that all primary and secondary event times are known (He et al., 2020). The implications of these approximations are also explored in Fig. 8.

##### 2.3.5 Interval-reduced censoring method

If we ignore the presence of right truncation, then Eq. (35) reduces to:

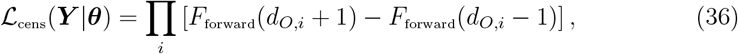

which, again, assumes a uniform distribution for *w*(*d*_*i*_). Reich et al. (2009) tested the consequences of this approximation and showed that assuming *w*(*d*_*i*_) is uniform results in low coverage probabilities (defined as the proportion of confidence intervals that contain the true value across multiple simulations). However, the uniform assumption remains in regular use in the applied literature.

##### 2.3.6 Truncation method

If we ignore the presence of censoring, Eq. (31) reduces to

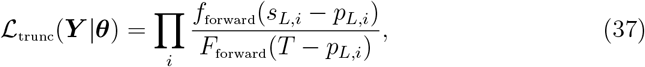

which can also be derived directly from Eq. (9) (Sun, 1995). Here, we use the lower bound for the primary event time to estimate the amount of truncation *T*− *p*_*L,i*_ as it represents the maximum degree of truncation. Other choices include using the upper bound *T*− *p*_*R,i*_ or the midpoint *T* − (*p*_*L,i*_ + *p*_*R,i*_)*/*2. The use of an upper bound may be numerically less stable because the upper bound of primary event time can be equal to the observation time, in which case the denominator becomes zero.

##### 2.3.7 Filtering method

A simple solution to mitigate truncation bias is to drop (or filter out) the most recent data (as these data will be most impacted by truncation) and then fit a probability distribution on the remaining observations. Given a filtering time *t*_filter_, the likelihood can be written as:

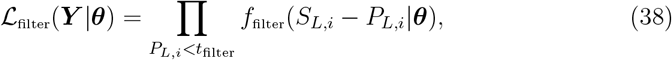

This approach can be readily extended to include double censoring as the filtering and distribution fitting steps are independent.

Unfortunately, whilst the filtering method may be convenient, it is sensitive to our choices of filtering time *t*_filter_. If *t*_filter_ is too early, we might not have enough data to estimate the focal delay distribution with certainty. If *t*_filter_ is too late, truncation bias might be introduced. More broadly, dropping existing data is often not a good practice and at the very least is likely to increase the uncertainty of any estimated distribution. This can be a particular issue during epidemics when data can be sparse and as accurate as possible estimates are needed to inform decision-making. For simplicity, we assume *t*_filter_ = 10 days throughout the paper.

##### 2.3.8 Naive method

A simple approach to estimating a delay distribution is to directly fit a probability distribution *f*_naive_ to the observed discrete delays. In this case, the likelihood is given by:

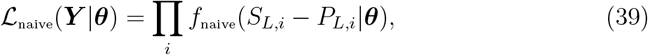

where ***θ*** represents the parameter vector and ***Y*** represents the data vector. This method does not account for truncation or censoring biases but was historically used in practice due to convenience and lack of awareness of its limitations. An even more basic form of this approach is to simply report the empirically observed summary statistics such as the mean, standard deviation, or range —this approach is common in practice (Nolen et al., 2016; Gostic et al., 2020; Miura et al., 2022). The no interval method in Fig. 2 highlights that this approach does not approximate the underlying generative process well in the presence of censoring.

##### 2.3.9 Discrete time methods for accounting for censoring and right truncation

So far, we have focused on fitting probability distributions to continuous forward delays directly by accounting for right truncation and censoring, presenting methods in order from more exact to most approximate. In this section, we outline discrete-time approaches to estimating epidemiological delay distributions whilst accounting for right truncation and censoring. These methods make the further simplifying assumption that censoring is daily for both primary and secondary events. To counterbalance this they both have a range of advantages to other methods in certain settings. In Fig. 9, we also present how different assumptions about the prior distributions within censoring intervals affect the discretisation of continuous-time distributions.

###### Dynamical correction method

In some cases, it may be favourable to exploit the relationship between backward and forward distributions to estimate the forward distribution. We refer to this method as the dynamical correction method because we explicitly take into account the biases in the backward distribution caused by epidemic dynamics. This approach may be particularly attractive when the backward epidemiological distribution has been reported without correction and the underlying data are not available or data on the time from primary event to observation time is not available. This is still common for incubation periods which are typically calculated by identifying symptomatic individuals and asking when they became infected.

Previously, we considered the distribution of the primary event time to account for the censoring problem in the forward distribution. Analogously, we now have to consider the distribution of the secondary event time to account for the censoring problem in the backward distribution. The probability that the primary event occurs between time *P*_*L*_ and *P*_*R*_ given that the secondary event occurred between time *S*_*L*_ and *S*_*R*_ can be then written as:

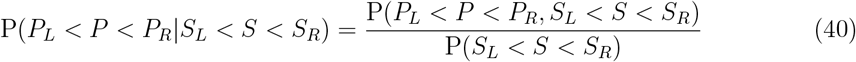

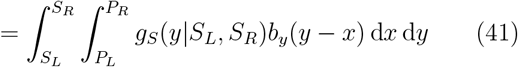

where *g*_*S*_(*y*|*S*_*L*_, *S*_*R*_) is the conditional probability distribution of the secondary event time given its lower *S*_*L*_ and upper *S*_*R*_ bounds. By substituting Eq. (4), we have:

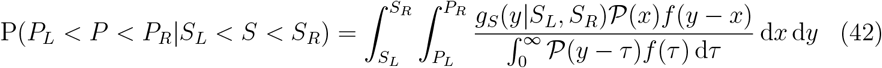

A special case of this framework is when the event times are exactly known and the incidence of primary events is changing exponentially at rate *r*. This special case was used by Verity et al. (2020a) to estimate the symptom-onset-to-death distribution of COVID-19 and has been previously introduced in several other contexts (Britton and Scalia Tomba, 2019; Park et al., 2021).

The general case presented in Eq. (42) depends on a continuous-time incidence pattern (Eq. (4)), but these incidence data usually only exist in discrete time. Therefore, given patterns of incidence of primary events reported on a daily basis 𝒫_*d*_(*t*), we approximate the continuous-time incidence as follows:

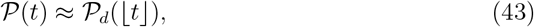

meaning that the primary incidence follows a step function, changing every day.

This approximation is convenient because it allows us to write the denominator of Eq. (42) in terms of a sum of integrals, which can be further simplified. In particular, setting *y* = *S*_*L*_ (i.e., the reported time of secondary event), we have:

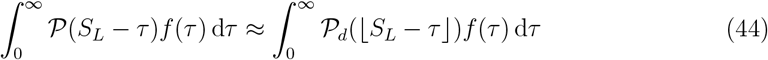

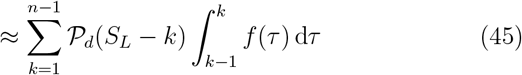

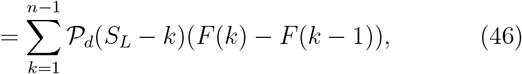

where *n* represents the length of the incidence time series. Likewise, the integral in the denominator can be further simplified:

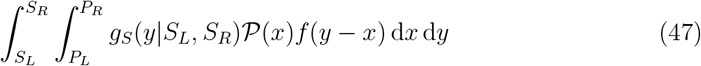

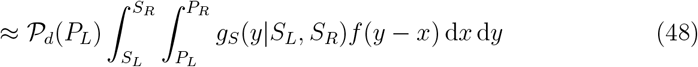

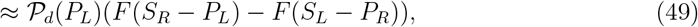

where the last line follows from the interval-reduced censoring approximation used previously (i.e., assuming a uniform censoring across the censoring window of the delay, rather than each event). Putting everything together, we have:

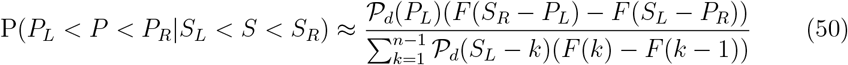

Finally, the likelihood for the model can be written as a product of the above probability:

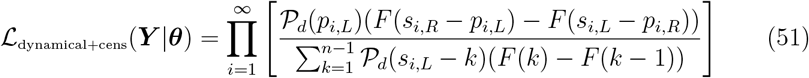

We note that these derivations rely on approximations that are specific to daily censoring and therefore will not necessarily hold for the general censoring case.

Unfortunately, this method requires a complete time series, or some approximation of it such as an estimate of the growth rate, in order to properly account for right truncation. In the following sections where we evaluate this approach, we first use a partially reported (i.e., truncated) time series of primary event incidence (directly aggregated from individual-level data) to evaluate the worst-case performance of this method for real-time usage. We also consider the best case where the complete incidence time series is known at the time of estimation. Approximations, such as using an estimate of the real-time growth rate or nowcast of primary incidence, would then perform intermediately between these two approaches depending on the accuracy of the approximation (i.e. the accuracy of the growth rate estimate or nowcast) and the propagation of uncertainty. Line lists may also not accurately reflect the incidence pattern when there are changes in surveillance and reporting and so the ability of this method to use potentially independent time series of incidence may be attractive.

###### Jointly modelling incidence and the forward distribution

Another alternative approach is to jointly model the discrete-time incidence of primary events and the forward delay from primary to secondary events as a count process. This method is more flexible than the previous dynamical approach as it does not require a complete time series to be available. It also allows for an error model to be applied to the primary incidence and can be naturally extended to include hazard effects and non-parametric delay distributions (Abbott et al., 2021) which may be desirable for complex, real-world, settings. This approach is routinely used in the nowcasting literature to reconstruct right truncated primary incidence curves (Abbott et al., 2021; Höhle and an der Heiden, 2014; Günther et al., 2021). A downside of this approach is that on top of specifying the reporting delay model, we must also define a model for the evolution of the expectation of primary incidence over time. Here we use a linear model for the exponential growth rate, though the package provides a large range of alternatives, to allow some flexibility in the primary event incidence time series.

As this method derives from the nowcasting literature it is commonly framed in terms of the “reporting triangle” (*p*_*t,d*_) (Höhle and an der Heiden, 2014). The reporting triangle defines the observed data as a matrix with the date of the primary event as the rows (*t*), and the date of the linked secondary event as the columns (*d*, relative to the time of the primary event). In contrast to previous methods, this frames the data as counts rather than as individual linked events. Primary event incidence 𝒫_*d*_(⌊*t*⌋) can then be recovered from this reporting triangle using

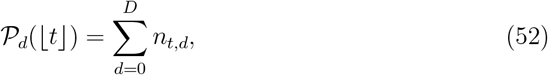

where *D* represents the maximum delay between the primary event and the secondary event which in theory could be infinite. However, in practice, we set it to a finite value in order to make the model identifiable and computationally feasible. For each *t*, events are assumed to be drawn from a multinomial distribution with *P*_*d*_(⌊*t*⌋) trials and a probability vector (*s*_*t,d*_) of length *D* indicating the probability of a secondary event after a given delay. We model this by estimating the components of this probability vector jointly with the expected number of primary events (*λ*_*t*_ = 𝔼[𝒫_*d*_⌊*t*⌋]) at time *t*. Here, we model the expected number of primary events (*λ*_*t*_) using an instantaneous daily growth rate model as follows,

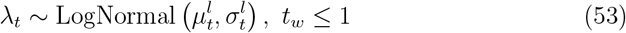

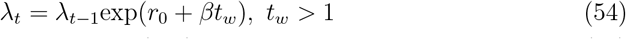

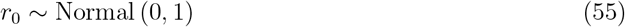

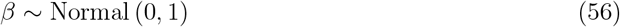

where the instantaneous growth rate (*r*_0_ + *βt*_*w*_) is defined using a linear model with *r*_0_ as the initial growth rate and *β* as the linear rate of change in the growth rate by week (*t*_*w*_). Note that this is an arbitrary model and that the precise specification should be appropriate to the modelled setting to avoid bias towards *T* (Lison et al., 2023). We assume that the delay distribution follows a daily discretised lognormal

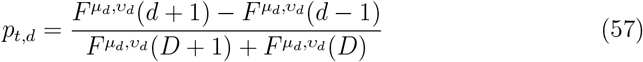

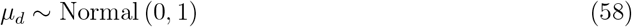

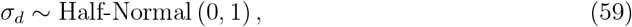

where 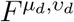 is the cumulative density function of the lognormal distribution, *μ*_*d*_ is the log mean, and *υ*_*d*_ is the log standard deviation. Note that this approach to calculating a probability mass function is equivalent to the interval-reduced method introduced above and discussed in Section 2.3.4 and Fig. 9.

Expected primary events (*n*_*t,d*_) by the time of primary event (*t*) and the time of the secondary event (*d*, relative to the time of the primary event) can now be calculated by multiplying expected primary events for each *t* with the probability of the secondary event occurring at a given date (*s*_*t,d*_). We assume a negative binomial observation model to account for potential overdispersion (with a standard half normal prior on 1 over the square root of the overdispersion (Stan Development Team, 2020)).

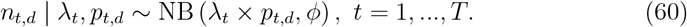

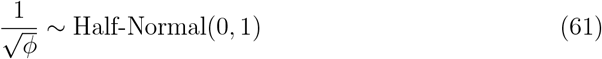

##### 2.3.10 Overview table

| Method | Details | Truncation | Censoring | Assumptions | Risk of bias |
| --- | --- | --- | --- | --- | --- |
| Exact latent variable censoring and truncation correction | Bayesian approach with the exact primary and secondary event times modeled as latent variables (using priors on their distribution over the censoring interval), and with a truncation likelihood integrating over the censoring uncertainty. | Yes | Yes | Exponential growth at a fixed rate within censoring interval. | Potential bias if growth rate changes quickly within the censoring interval. |
| Approximate latent variable censoring and truncation correction | Approximation of exact latent variable approach that results in a simpler likelihood identical to the plain right truncation case (no integration over primary event times required). Censoring uncertainty is only expressed via priors on the latent even time variables. | Yes | Yes | Primary event time is uniformly distributed between the lower and upper bound, and censoring interval of primary events is sufficiently narrow for the cumulative delay distribution to be approximately constant within its window. | Biased if the primary event times are not uniformly distributed. Small bias from the constant approximation of cumulative delay distribution within the censoring interval. |
| Interval-reduced censoring and truncation correction | Reformulation of the censoring and truncation likelihood that integrates over the delay (which is a convolution of the primary and secondary event time conditional distributions). Additional assumptions lead to an approximation, which does not require any integration. | Yes | Yes | Delay between primary and secondary events is uniformly distributed within its lower and upper bounds, and censoring interval of primary events is sufficiently narrow for the cumulative delay distribution to be approximately constant within its window. | The assumption of uniformly distributed delays within their bounds is likely unrealistic and deviates from the delay distribution that would result from uniformly distributed event times. Small bias from the constant approximation of cumulative delay distribution within a censoring interval. |
| Interval-reduced censoring correction | Same likelihood as interval-reduced censoring and truncation, but without conditioning on observation up to present. | No | Yes | Same as interval-reduced censoring and truncation, but additionally assuming no right truncation, i.e. all relevant event pairs have been observed. | Right truncation bias (leading to underestimation of mean) in addition to the biases described above. |
| Truncation correction | Likelihood for delay distribution that uses lower bounds of event times (or different point in censoring interval, e.g. midpoint), conditioned on observation up to present. | Yes | No | Assumes no censoring, i.e. that true event times match the assumed event times within censoring interval. | Censoring bias (leading to over-estimation of standard deviation) |
| Filtering | Exclusion from the likelihood of observations with primary event after some chosen filtering time, and likelihood for delay distribution that uses lower bounds of event times (or different point in censoring interval, e.g. midpoint). | Yes | No | Assumes no censoring, i.e. that true event times match the assumed event times within censoring interval. | Truncation bias if filtering time is too late, censoring bias. |
| Filtering + censoring correction | Same as filtering, but with a likelihood that accounts for interval-reduced censoring. | Yes | Yes | Assuming interval-reduced censoring (i.e., uniformly distributed delay within the upper and lower bounds). | Truncation bias if filtering time is too late and biases from interval-reduced censoring. |
| Naïve estimation | Likelihood for delay distribution that uses lower bounds of event times (or different point in censoring interval, e.g. midpoint), not conditioned on observation up to present. | No | No | Assumes no right truncation, i.e. all relevant event pairs have been observed, and no censoring, i.e. that true event times match the assumed event times within censoring interval. | Both right truncation bias and censoring bias. |
| Dynamical censoring correction (discrete) | Likelihood motivated from modeling the backward delay distribution under epidemic dynamics, tailored to the case of a discrete incidence time series. | Partially (depending on assumed incidence time series) | Yes | Discretisation of continuous incidence to dates (step function), centered on the lower bounds of primary and secondary events, respectively, and delay between primary and secondary event is uniformly distributed. | The assumption of uniformly distributed delays is likely unrealistic and deviates from the delay distribution that would result from uniformly distributed censoring times. Biased if the estimate of incidence time series is biased. Can also have a small discretisation bias. |
| Joint incidence and forward delay estimation (discrete) | Latent-variable nowcasting model, using a count likelihood for case counts by primary event and delay, with an epidemiological prior on the case counts by primary event (e.g. exponential growth model), and interval-reduced censoring of the delays. | Yes | Yes | Assumes that case counts in the cells of the reporting triangle have a certain count distribution, that incidence follows a certain time series model, and that delay between primary and secondary event is uniformly distributed. | The assumption of uniformly distributed delays is likely unrealistic and deviates from the delay distribution that would result from uniformly distributed censoring times. Potential bias if the epidemiological model for incidence is inappropriate. |

### 2.4 Simulation study

In order to understand the impact of different biases and the performance of the various methods that we have outlined, we conducted a series of simulation studies designed to replicate different real-world scenarios.

#### Summary

- We evaluated the methods discussed above across a range of simulated exponential growth and epidemic wave scenarios.
- To simulate exponential growth settings, we used a continuous-time individual-based exponential growth model and to simulate epidemic wave settings, we used a stochastic Susceptible-Infected-Recovered model.
- As this study was initially inspired by the analysis of COVID-19 outbreaks, we considered settings with a range of delay distributions to assess the impact of common biases across a range of common scenarios for COVID-19. These delays had the following means (and standard deviations) on the natural scale:
- 3.6 (1.5) days, 5.9 (3.9) days, and 8.3 (7.9) days. We refer to these in the text as “short”, “medium”, and “long” delays.

##### Simulating infections

We explored both fixed growth rate and epidemic scenarios. For all scenarios, we explored 3 lognormal distributions representing “short” (log mean of 1.2 and log standard deviation of 0.4), “medium” (log mean of 1.6 and log standard deviation of 0.6), and “long” (log mean of 1.8 and log standard deviation of 0.8) time delays. These assumptions correspond to distributions with the following means (and standard deviations) on the natural scale: 3.6 (1.5) days, 5.9 (3.9) days, and 8.3 (7.9) days (Fig. 4B).

To simulate settings with a fixed growth rate, we used continuous-time stochastic simulations of exponential growth with a range of daily growth rate assumptions (assuming 10,000 individuals): “fast decay” (−0.2), “decay” (−0.1), “stable” (0), “growth” (0.1), and “fast growth” (0.2).

To simulate epidemic scenarios, we used a stochastic Susceptible-Infected-Recovered (SIR) model implemented using a Gillespie algorithm assuming an early exponential growth rate of 0.2 per day and a recovery rate of 1/7 per day (corresponding to a mean of 7 days) with 50 initial cases and a total population of 10,000.

For all scenarios, we took the primary event time to be the time of infection, and we simulated secondary events by drawing delays from the assumed continuous lognormal distribution for that scenario for each individual and then adding this delay to their primary event time. This simulation approach assumes no observation error for the primary event or secondary events beyond daily censoring and thus represents an idealised system.

##### Scenarios investigated

For the simulations with fixed growth rates, we assumed a sample size of 200 randomly drawn pairs of primary and secondary events with observation cut-off 30 days after the start of the simulation. We repeated each sampling step 20 times independently so that we ended up with 20 replicates of each simulated growth rate scenario.

For the epidemic wave simulation, we explored a range of sample sizes (10, 100, 200, and 400 samples) and observation time scenarios (including all observations up to 15 days, 30 days, 45 days, and 60 days from the start of the simulation).

### 2.5 Case study: 2014-2016 Sierra Leone Ebola virus disease epidemic

In order to explore the performance of each method in a real-world setting, we used publicly available data from the 2014-2016 Sierra Leone Ebola virus disease epidemic.

#### Summary

- We studied the evolution over time of the empirically observed backward and forward delay distributions of the time from symptom onset to positive test, and the proportion of positive tests that were unobserved for a cohort of symptom onsets over a rolling 60-day observation period.
- We used multiple observation times and compared retrospective estimates to real-time estimates for each approach.

##### Data

We downloaded line list data from Fang et al. (2016) which contained age, sex, symptom onset date, sample test date, the district of the case, and the Chiefdom of the case for Ebola virus disease (EVD) cases in Sierra Leone from May 2014 through September 2015. We then processed these data to keep only the date of symptom onset and the date of the sample test. We assumed that censoring for each of these dates was daily with a day defined as being from 12:00 AM to 11:59 PM.

##### Empirical context

We calculated various summary statistics using the available line list data, which we used to provide a context for comparisons of different inference methods. These statistics included changes in the mean forward and backward distributions, the empirical forward distribution at each observation cut-off date, as outlined in the ‘Scenarios Investigated’ section, in both real-time and retrospective settings; and, the proportion of secondary events that were unobserved for a cohort of individuals whose primary events took place within a rolling 60-day window.

##### Scenarios investigated

We estimated the delay from symptom onset to sample test using each method across four different observation windows of 60 days each (0–60 days, 60–120 days, 120–180 days, and 180–240 days after the first symptom onset). In particular, for each window, we only considered individuals who developed symptoms and also got tested within the time period to match a real-time analysis setting. Note that in this case study we do not account for additional potential delays from the sample test date to the reporting of these tests as these data were not available in Fang et al. (2016). However, our methods could naturally account for these additional delays if data were available. As a comparison, we then re-estimated the delay using each method for each observation period by including all individuals who developed symptoms within the observation period (regardless of when they got tested) and estimated the delay distribution. These retrospective estimates were then used to represent the “true” distribution when calculating the relative difference between estimates using data available in real-time and data available retrospectively for each method. Note that in this setting the “true” distribution may still be biased relative to the underlying forward distribution (e.g. methods that do not account for censoring may still be biased in both real-time and retrospective settings).

### 2.6 Evaluation

We evaluated the recovery of the mean and standard deviation of the lognormal distribution in our simulated scenarios visually using the posterior density normalised with known synthetic values, where these were available, and quantitatively using the Continuous Ranked Probability Score (CRPS, Gneiting and Raftery, 2007), which is a generalisation of absolute error. The CRPS measures the distance between a distribution and data as follows

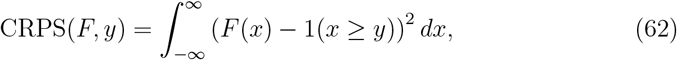

where *y* is the true observed value and *F* is the cumulative distribution function (CDF) of the predictive distribution, and 1(*x* ≥ *y*) is the indicator function such that for *x* ≥ *y* its value is 1 and otherwise it is 0. The CRPS is a strictly proper scoring rule, meaning that only a probabilistic estimate that exactly reflects the true distribution of *y* minimises the expected score.

To allow for comparisons across simulated scenarios, we normalised predictions by the known true value, took the natural log, and then calculated the CRPS. In effect, this transforms the CRPS into a relative, rather than an absolute, score whilst maintaining its propriety (Bosse et al., 2023). We then averaged across all observations, and then averaged across observation stratified by growth rate, parameter, and delay distribution.

As a comparison, we also compute relative bias, root mean squared error (RMSE), and coverage probabilities. The relative bias and RMSE were calculated using the log of relative errors (i.e., the ratio between the estimated and true values) in the same way as the CRPS. The coverage probability was calculated from the proportion of 90% credible intervals that contain the true value.

### 2.7 Implementation

All models were implemented using the `brms` (Bürkner, 2018) R package and the `stan` probabilistic programming language (Stan Development Team, 2021) (Gabry and Češnovar, 2021) in `R` version 4.2.2 (R Core Team, 2019). As such they are readily extensible to model formulations covered by the GAM framework. Where not otherwise specified, we use the following priors for all methods,

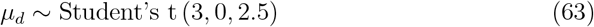

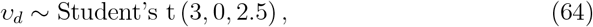

where *μ*_*d*_ is the log mean, and *υ*_*d*_ is the log standard deviation of the lognormal distribution. Here, the student’s t distribution has the following three parameters: degrees of freedom, mean, and standard deviations.

The No-U-Turn Sampler (NUTS), which is an adaptive variant of Hamiltonian Monte Carlo (HMC) was used for model fitting via `cmdstanr`. We used four Markov chain Monte Carlo (MCMC) chains with each having 1000 warm-up and 1000 sampling steps (Gabry and Češnovar, 2021); we did not consider any other sampling schemes or settings for simplicity. We set `adapt delta`, which represents the target average acceptance probability, to 0.95 due to the expected complexity of the posterior distribution (Betancourt, 2017).

We assessed convergence using the Rhat diagnostic, where an R-hat close to 1 indicates that the MCMC chains have converged (we used 1.05 as a threshold for further investigation) (Gelman and Rubin, 1992), and recorded run-time, the number of divergent transitions, the exceedance of the maximum tree depth (which was 10, its default setting), and the effective sample size (Gabry and Češnovar, 2021; Stan

Development Team, 2021). We did not assess the sensitivity of our inference to these settings. Divergent transitions are useful as a model diagnostic when using HMC since they indicate issues exploring the posterior, and when present, can mean that estimates are unreliable (Betancourt, 2017).

The `scoringutils` R package (1.1.0) (Bosse et al., 2022) was used to calculate the CRPS, RMSE, and to assess bias. We also evaluated the sampling performance details of each method, including the run-time of inference and the distribution of divergent transitions.

All core functionalities and inference approaches were implemented in the `epidist` R package to facilitate reuse and user extension. Our simulation, scenario, and model fitting pipeline was implemented in a `targets` workflow (Landau, 2021) and we provide an archive of our results to aid reuse. Our post-processing pipeline was also implemented as a `targets` workflow.

To enhance the reproducibility of this analysis, we managed dependencies using the `renv` R package (Ushey, 2021). In addition, we provide a versioned Dockerfile and a prebuilt archived image (Boettiger, 2015). The code for this analysis can be found here: https://github.com/parksw3/epidist-paper. The code for the `epidist` R package can be found here: https://github.com/epinowcast/epidist.

## 3 Results

### 3.1 Simulation study

#### Summary

- Not accounting for the truncation bias led to an underestimation of the mean. The degree of bias increased with the underlying growth rate of an epidemic.
- Not accounting for the censoring bias generally led to over-estimation of the standard deviation. For daily reporting, assuming uniform censoring for each event gave reasonable answers (i.e., the interval-reduced approach).
- Among methods that explicitly account for the censoring and truncation bias in the forward distribution, the approximate-latent-variable approach performed the best overall, followed by the interval-reduced-censoring-and-truncation approach.
- The joint modeling and dynamical correction methods had nearly comparable performance. However, the joint modeling approach had wider uncertainty than comparable approaches whilst the dynamical correction method was unreliable during negative growth periods and required a retrospective time series to perform well.

#### 3.1.1 Exponential growth simulation

The joint modelling approach performed best overall when evaluated using relative CRPS (Fig. 3C), followed by the approximate-latent-variable model, the interval-reduced censoring method, and the retrospective dynamical correction approach. All of these approaches successfully recovered the simulated parameters to a reasonable degree both visually (Fig. 3C), based on both probabilistic and point scores (Fig. 10A), and based on coverage of the 90% credible interval (Fig. 10C). These methods performed worse for long delays with both an increase in uncertainty and in underestimation of both the mean and standard deviation. These biases were a particular issue for higher growth rates. The approximate-latent-variable model appeared most robust to this bias visually, though this was not conclusively demonstrated by any of the evaluation metrics used. The dynamical correction method using retrospective incidence data was the least robust of the methods that performed well when growth rates were negative with biased estimates of the standard deviation across all delay distributions explored. Unlike other well-performing methods, the joint modelling approach had less variance in its relative CRPRs scores without the very low scores for large growth rates and small delays scenarios but also without reduced performance for scenarios with longer delays for the mean (though not the standard deviation).

**Figure 3:**
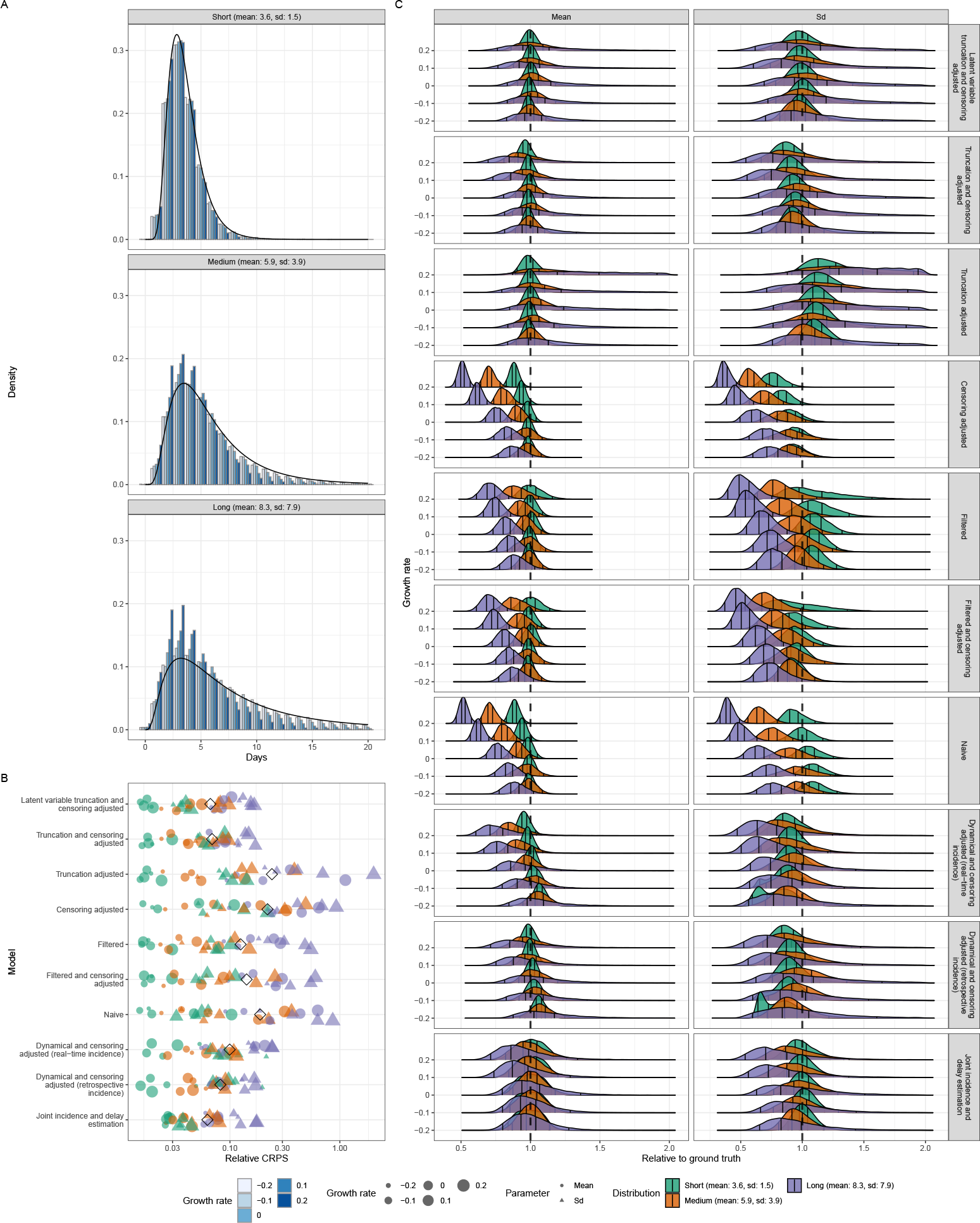
Simulation study with fixed growth rates. (A) Empirical distribution of the delay between primary and secondary events stratified by distribution. Shade indicates the exponential growth rate used in each simulated scenario. The black line indicates the true distribution used for simulation. (B) Relative CRPS of each method (smaller is better). Here black diamonds represent the global relative CRPS. Coloured points represent the mean CRPS for each distribution scenario (short, medium, and long) with the shape indicating if the score is for the mean (circle) or standard deviation (SD, triangle). (C) Posterior distributions of mean and standard deviation (Sd) relative to the values used when simulating. The vertical line at 1 indicates exact replication of the true value. Vertical lines represent the 5%, 35%, 65%, and 95% quantiles respectively. Models are ordered based on the order used in the methods section.

The dynamical correction method with real-time incidence performed better than other methods that do not fully account for censoring and truncation. However, it performed worse than all other methods that account for these biases. In the real-time setting, the dynamical correction method had similar characteristics as when used with retrospective data, but showed more under- (when growth rates were positive) and over- (when growth rates were negative) estimation of the mean due to the use of truncated real-time incidence. Real-world performance for this approach would be somewhere between the real-time and retrospective approaches, depending on the availability of robust estimates for primary incidence that correct for truncation (or alternatively a robust estimate of the growth rate in stable growth settings).

All other models perform considerably worse than these methods with methods that only accounted for either truncation or censoring performing the worst. The naive method that accounts for no biases outperformed these methods as censoring and truncation biases partially cancelled each other when growth was fast (as the direction of these mechanisms bias estimates in different directions). Out of the poorly performing methods, the filtering methods performed the best with the potential to perform even better if the filtering horizon were optimised to the length of the delay (though at the cost of increased uncertainty). All of these methods under-covered, indicating that their credible intervals were too narrow, and produced biased estimates (Fig. 10).

Methods that did not adjust for right truncation produced increasingly underestimated means and standard deviations as the exponential growth rate increased (*r >* 0) with the degree of underestimation increasing with the length of the delay distribution. Whilst methods that do explicitly account for the truncation bias generally give unbiased estimates of the mean (Fig. 3C) when the growth rate is high and the delay is longer (i.e., when the degree of truncation is large), the truncation method without censoring overestimated the mean and standard deviation. Better performing methods also struggled in this extreme setting with larger credible intervals for the standard deviation though the estimates were visually still close to the true values.

#### 3.1.2 Epidemic wave simulation

For the epidemic wave simulation, the ordering of well-performing methods differed from that of the exponential growth simulations but the performance difference between methods that did and did not account for both censoring and right truncation remained. Here the approximate-latent-variable approach performed the best overall (Fig. 4C) based on relative CRPS, followed by the dynamical approach using retrospective incidence, the joint modeling approach, and finally the interval-reduced-censoring-and-truncation approach.

**Figure 4:**
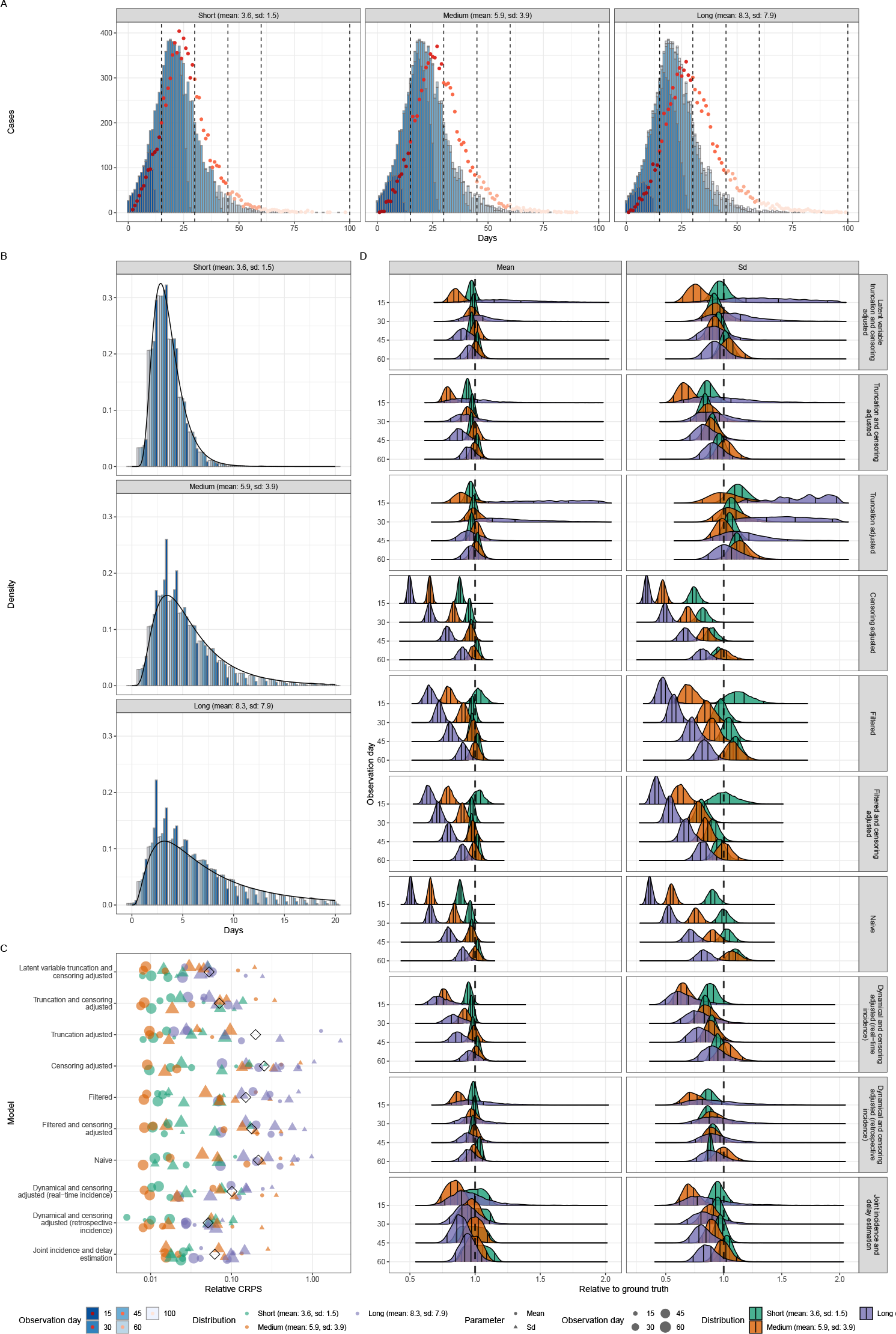
(A) Epidemic wave simulations across three distributions. Dashed bars represent four observation days. Blue bars represent the observed incidence of primary events on each observation day. Red points represent the observed incidence of secondary events on each observation day. On each observation day, we take 400 random samples from simulated primary and secondary event pairs and fit lognormal distributions while accounting for truncation and censoring biases. (B) Empirical distribution of the delay between primary and secondary events stratified by distribution. Shade indicates the day of observation. The black line indicates the true distribution used for simulation. (C) Relative CRPS of each method (smaller is better). Here black diamonds represent the global relative CRPS. Coloured points represent the mean CRPS for each distribution scenario with the shape indicating if the score is for the mean (circle) or standard deviation (Sd, triangle). (D) Posterior distributions of mean and standard deviation (Sd) relative to the values used when simulating. The vertical line at 1 indicates exact replication of the true value. Vertical lines represent the 5%, 35%, 65%, and 95% quantiles respectively. Models are ordered based on the order used in the methods section.

Early in the epidemic, all of these methods perform worse when combined with longer delays. This setting was particularly problematic for the approximate-latent-variable and the interval-reduced-censoring-and-truncation approaches (Fig. 4D), giving wider uncertainty intervals. Underestimation of the standard deviation was more common in the epidemic wave simulations across observation windows than it was in the exponential growth scenarios for these methods, with the approximatelatent-variable method being the least impacted.

As in the exponential growth simulations, the dynamical correction approach with real-time incidence performed less well than other methods that sought to account for both right truncation and censoring due to the use of a truncated time series. Methods that did not account for both censoring and truncation also give biased estimates, especially at the beginning of the epidemic (i.e. when exponential growth was highest).

### 3.2 Case study: 2014–2016 Sierra Leone Ebola virus disease epidemic

#### Summary

- The epidemic had four distinct phases: sporadic cases in the first phase, rapid growth in the second phase, plateau in the third phase, and decay in the fourth phase.
- The delay in reporting cases (from symptom onset) increased as the number of cases grew, reaching a maximum mean delay of around 7 days in the third phase and then declining, potentially reflecting changes in testing capacity, reporting, or other mechanisms linked to incidence.
- The forward and backward delay distributions exhibited similar trends, but with differences in mean delay during the growth and decay periods. Right truncation during the period of faster growth resulted in a difference of approximately 0.5 days in the mean delay between real-time and retrospective observations. The degree of right truncation was relatively small due to the growth rate of this epidemic also being relatively small even during the peak growth period.
- Methods that considered right truncation and censoring performed well, producing real-time estimates that closely matched retrospective estimates, similar to the findings in the simulation studies.
- Differences between real-time and retrospective estimates were largest during the period of exponential growth. All well-performing methods slightly overestimated the retrospective standard deviation in real-time during this period.
- The joint modeling approach had larger credible intervals compared to other methods that accounted for right truncation and censoring and also slightly overestimated the retrospective mean in real-time, this was observed to some degree in other methods to a lesser extent, except for the dynamic correction method using retrospective incidence data.

#### 3.2.1 Empirical observations

During the first phase of the epidemic, cases were reported sporadically without apparent growth (Fig. 5A). The epidemic then grew rapidly during the second phase, followed by a plateau during the third phase and decay during the fourth phase. For each phase, we used all available samples from the last 60 days which resulted in real-time (and retrospective) sample sizes of 834 (1032) at 60 days, 3149 (3532) at 120 days, 2399 (2483) at 180 days, and 401 (428) at 240 days.

**Figure 5:**
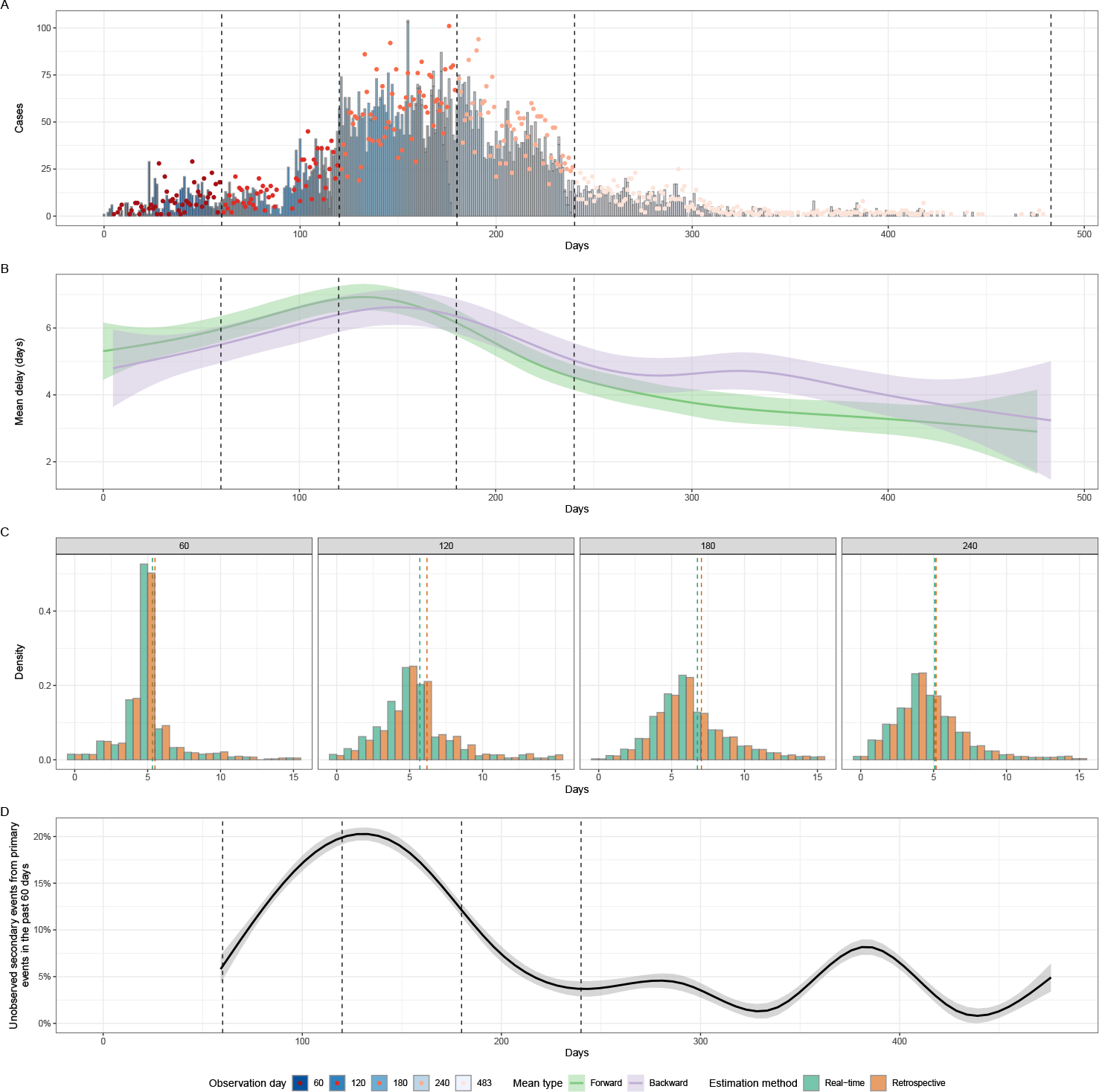
(A) Reported number of Ebola virus disease cases between May 18, 2014 and September 13, 2015. Dashed lines represent four observation days. Blue bars represent the daily incidence of primary events with different shades indicating the observed incidence in each observation period. Red points represent the daily incidence of secondary events with different shades indicating the observed incidence in each observation period. (B) Mean forward (measured across a cohort of individuals who developed symptoms on the same day) and backward (measured across a cohort of individuals who reported their infections on the same day) delays between symptom onset and case reports and the corresponding 95% confidence intervals. A Generalised Additive Model (GAM) with a default thin plate spline on the date of the event has been used to smooth daily estimates. (C) Empirical distribution of the delay between primary and secondary events both in real-time (green) and retrospectively (orange) stratified by observation. The dotted lines indicate the observed mean both in real-time (green) and retrospectively (orange). (D) Proportion of unobserved secondary events (and therefore, delays) from primary events in the last 60 days. A GAM with a default thin plate spline on the date of the primary event has been used to smooth daily estimates with the point estimate and its 95% confidence interval shown.

The forward delay distribution (as observed retrospectively) changed over the course of the epidemic (Fig. 5B), potentially reflecting changes in the reporting process. During the initial phase, the delays were short with means around 5 days. As the number of cases increased, the mean delay also started to increase, potentially reflecting an overload in a testing capacity or another mechanism linked to incidence. During the third phase, the mean delay reached its maximum of around 7 days and started to decline along with incidence. Eventually, the mean delay decreased to 5.9 days by day 240. The backward distribution exhibited a similar trend, though as explained earlier, it had a shorter mean than the forward distribution during the growth period and a longer mean during the decay period.

The empirical distributions for each observation period, both as observed in realtime and retrospectively, are presented in Fig. 5C. Delay distributions observed in real-time (truncated) and retrospectively (untruncated) were broadly similar across all observation windows, except for days 60–120 during the period of rapid growth in incidence. During this period, the difference in the mean delay was around 0.5 days with this difference reflecting large amounts of right truncation (Fig. 5D).

#### 3.2.2 Model results

The joint modelling approach, the approximate-latent-variable model, the interval-reduced censoring method, and the retrospective dynamical correction approach again performed well in real-time, producing estimates that were comparable to those estimated using retrospective data (Fig. 6A, Fig. 12A). As expected, the largest differences between retrospective and real-time estimates are observed during the period of exponential growth (i.e., 60-120 days). During this period, all of these methods’ real-time estimates overestimated the retrospective estimates of the standard deviation. The joint modeling approach had significantly wider credible intervals across all observation periods compared to other well-performing methods. The joint modeling approach also routinely overestimated the retrospective mean in real-time settings. We note that the same overestimation occurred to some degree for all other methods aside from the dynamical correction method using retrospective incidence data.

**Figure 6:**
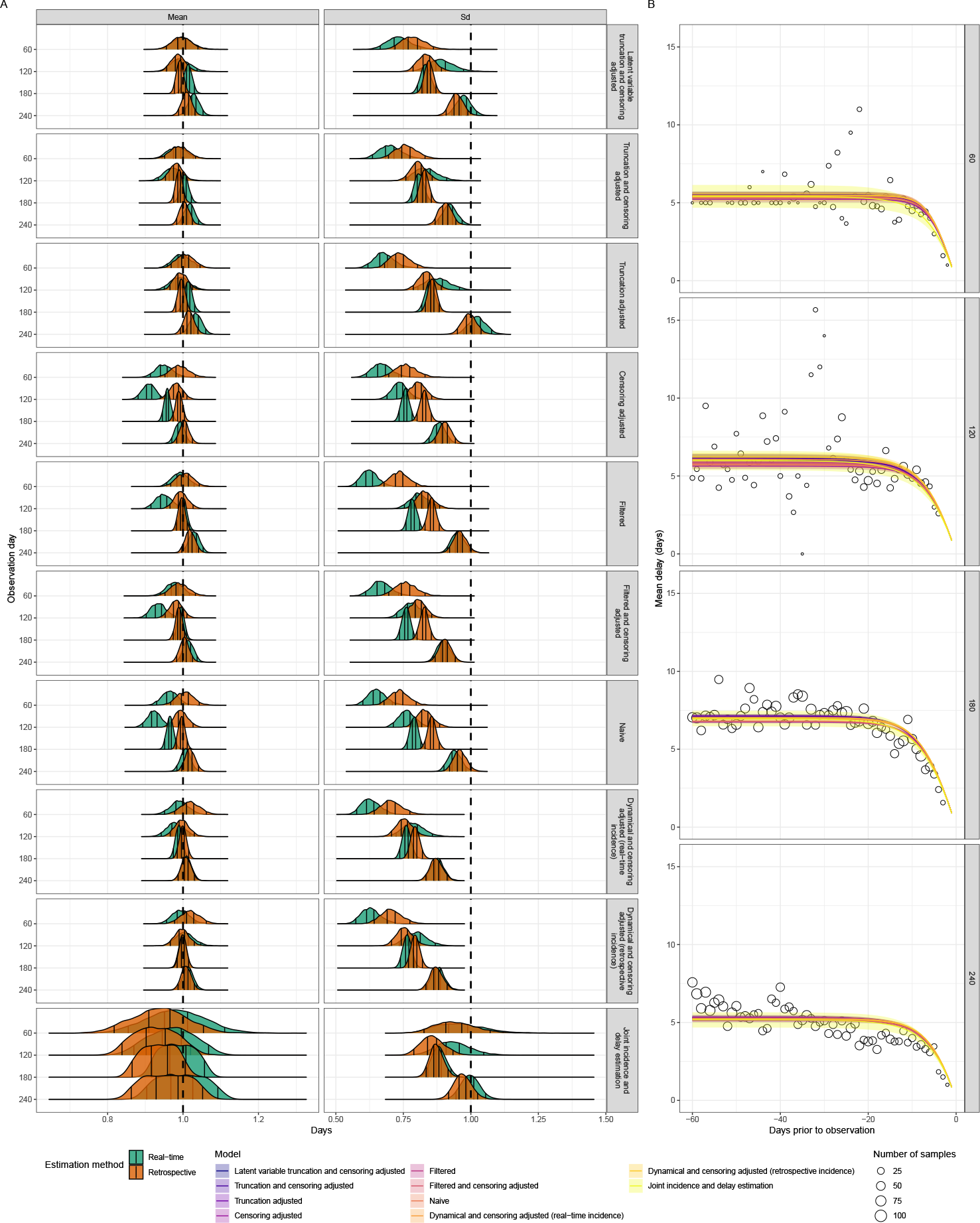
(A) Posterior distributions of mean and standard deviation (Sd) normalised by the retrospectively observed mean and standard deviation from the data. Vertical lines represent the 5%, 35%, 65%, and 95% quantiles respectively. Dashed vertical lines correspond to the unbiased estimate. Models are ordered based on the order used in the methods section. Recovery of the empirical standard deviation is not expected as the empirical values are biased due to the censoring process. (B) Posterior predictions of the truncated mean over time compared to the observed forward mean. Lines and shaded regions represent posterior median and 95% credible intervals. Points represent the observed mean values.

These well-performing methods also generally captured the retrospective empirically observed mean well. However, the estimated standard deviations are considerably lower than the empirical values (Fig. 6A). While these differences likely reflect the bias in the empirical values caused by the censoring process, it is also possible that the lognormal distribution may not be the best choice of distribution for these data. The interval-reduced and dynamic correction methods particularly underestimated the empirically observed standard deviation.

As in our simulation scenarios, not accounting for truncation gave real-time fits with lower estimates of the mean and standard deviation than the corresponding retrospective fit (Fig. 6A, Fig. 12A). As both retrospective and real-time fits are liable to censoring bias, the impact of not properly accounting for censoring is not highlighted by this case study. This also means that the estimates for the standard deviation from the truncation-only adjusted model should best reflect the empirical standard deviation, as both are biased upwards due to censoring when compared to the standard deviation of the continuous distributions.

All methods were able to reasonably match the empirical mean of the data in each observation period despite assuming a constant mean and standard deviation within each period (Fig. 6B). Due to the relatively slow rate of exponential growth for all observation periods, the absolute differences between the estimated means for most methods were relatively small.

#### 3.2.3 Implementation considerations

##### Summary

- More complex methods for estimating epidemiological delay distributions have increased computational requirements relative to simpler methods. In most settings, these computational requirements are expected to still be feasible for routine usage.
- The approximate-latent-variable method had exponential scaling in resource requirements when the sample size increased.
- The interval-reduced-censoring-and-truncation method required the least computational resources by an order of magnitude of the methods that accounted for both censoring and truncation.
- The dynamical adjustment method is unstable for short delays with larger sample sizes both with and without a truncated time series, though the instability increased when a truncated time series was used (i.e., a real-time one).

All the methods we considered are implementable with modest (i.e., laptop scale) computational hardware at the time of writing, and we are able to run our full analysis pipeline of several thousand model fits within this resource budget at a practical time scale (i.e, within several hours). However, more complex methods required greater computational resources. For example, the approximate latent variable, dynamic correction, and joint modeling approaches required an order of magnitude more resources for the same sample size than the interval-reduced-censoring-and-truncation method (Fig. 7A). The resource requirements for all models scaled with sample size, with the approximate latent variable model scaling the worst of the methods we explored. The dynamical correction approach had the highest variance in its computational requirements with some fits taking 10 to 100 times longer despite having the data used having the same sample size across fits. This was likely due to numerical instability from the integration step. It was a particular issue when a real-time incidence time series was used.

**Figure 7:**
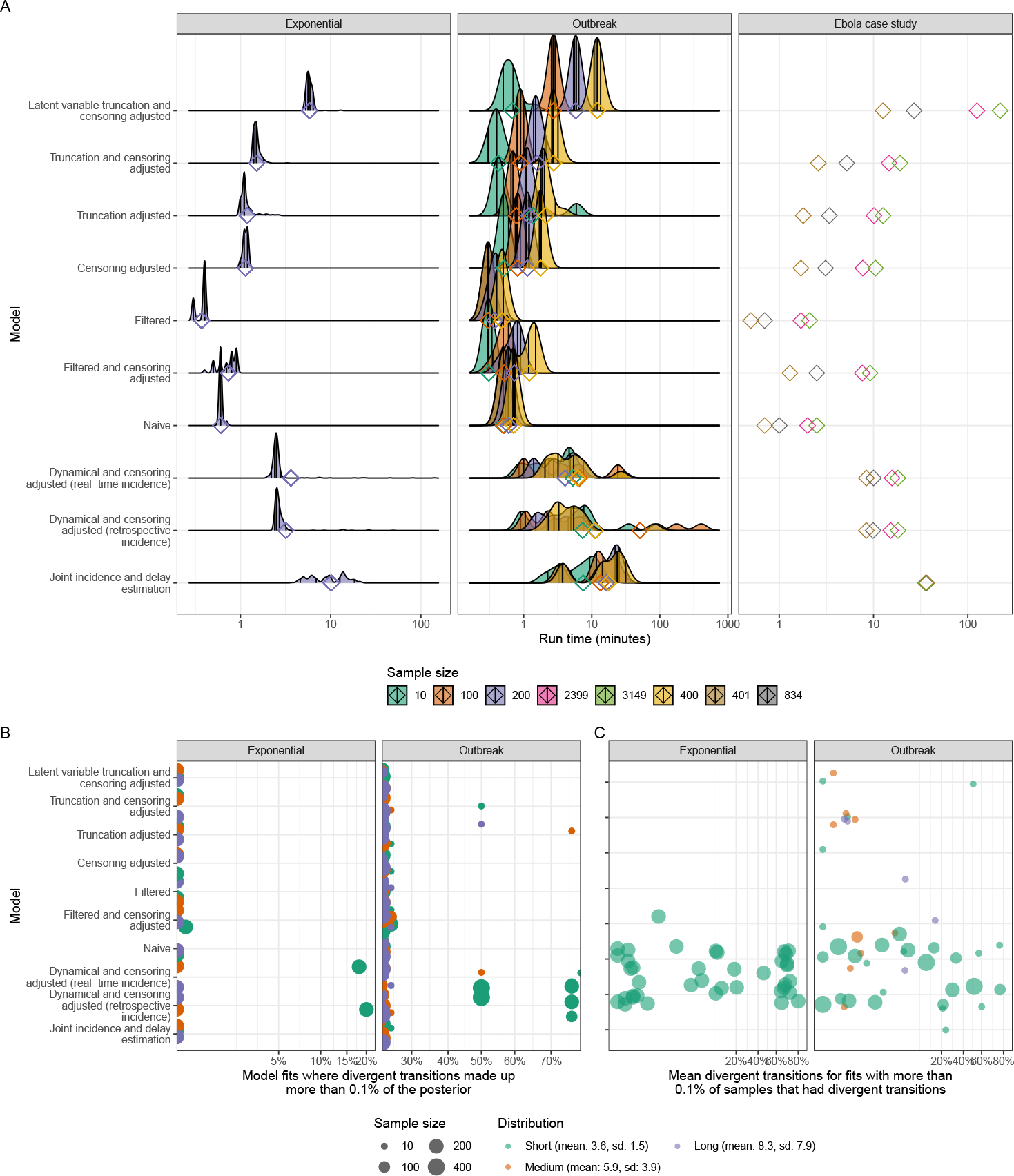
(A) Distributions of run-times for model fitting using a 2019 16-core AMD Threadripper on the log 10 scale. Diamonds represent the overall mean run times and vertical lines represent the 5%, 35%, 65%, and 95% quantiles respectively. Whilst run times are specific to the hardware used, the relative differences between models and scenarios should be more readily generalisable to other hardware. (B) Percentage of model fits with more than 0.1% divergent transitions on the logit scale. (C) Percentage of divergent transitions for fits with more than 0.1% of samples that had divergent transitions on the logit scale.

In general, all the methods were numerically stable, excluding the dynamical correction approach in some settings, and converged. Methods that better captured the data-generating process (i.e. that accounted for known biases) were the most stable and had fewer diagnostic warnings. A notable exception to this was the dynamical model which was the only model to cause sampling in `stan` to fail completely in six instances and more generally had the highest proportion of fits with divergent transitions (Fig. 7B). These issues occurred most frequently in simulations with short delays and during periods of epidemic decline. In the case study, both retrospective and real-time fits failed at the 180-day observation point. Another driver of model issues was low sample sizes with the majority of instances of divergent transitions occurring in settings with a sample size of 10.

## 4 Discussion

### 4.1 Findings

In this work, we provided methodological and practical guidance for researchers tasked with estimating epidemiological delay distributions. We first introduced the general theory of epidemiological delay distributions and the most common kinds of biases (namely censoring, truncation, and dynamic) that can impact their inference. Based on this theory, we derived an exact approach for accounting for these biases when estimating epidemiological delay distributions. As this approach lacks stability and practicality for real-time usage, we then presented a set of methods that approximate the exact solution, and compared their performance. We made use of simulated scenarios and a case study using data from the 2014-2016 Sierra Leone Ebola virus disease epidemic to compare these methods in applied usage, evaluating not only their accuracy and calibration but also practical issues, such as their suitability for real-time usage, their computational requirements, and their numerical stability.

We showed that naive methods that correct for none or only one form of bias can severely misestimate the distribution mean, e.g. by up to 50% in the most extreme cases we studied. Generally, we suggest using the approximate latent variable model Ward et al. (2022), which explicitly adjusts for both censoring and truncation biases, and can produce much more accurate estimates both in real-time and retrospectively. However, this method was not foolproof and could not estimate the distribution mean or standard deviation with precision when the exponential growth rate was very high and the true delay distribution was long. If this method is computationally too costly or complex to implement, using the interval-reduced censoring and truncation model (which assumes a 2-day censoring window around each observed delay) gives only slightly more biased estimates. The joint modeling approach and dynamic correction methods also performed well (in the latter case only when a retrospective time series was available) and may be sensible choices in some settings. In the case of the joint model, these settings are likely to be those in which a nowcast of the primary event is useful, where the primary event has observation error, or when a hazard-based approach would allow additional complexities in the reporting process to be modelled. However, this model’s requirement for a primary incidence model to be specified may make it more complex to use more generally. Similarly, the dynamical correction approach’s requirement for an untruncated time series generally means another model would first need to be used to estimate the incidence time series. This process is likely to introduce hard-to-quantify bias and make propagating uncertainty difficult (in the absence of a similar joint approach we have explored for the forward distribution). However, where an independent time series is available that does not suffer from right truncation this approach would be more practical. Finally, we provide all the methods we have evaluated as a standalone, and readily extensible, `R` package (`epidist`), which additionally provides functionality to fit distributions that are partially pooled and that vary in discrete time.

Right-truncation bias was most pronounced for growing epidemics and long delays. Failing to account for this bias typically led to an underestimation of the mean. This underestimation increased with higher degrees of truncation. We have discussed several methods that can robustly adjust for right truncation, but when the degree of right truncation was very large, even these well-performing methods overestimated the mean. Though their credible intervals still covered the true mean in all cases, performance could likely be improved by using a more informed prior distribution. For this reason, it may be helpful for practitioners to track the degree of expected right truncation in real time. Unfortunately, this was difficult to do, but one heuristic approach is to plot the forward mean (Fig. 5B) as well as the proportion truncated (Fig. 5D) over time and compare their changes. Comparing retrospective estimates with real-time estimates is also helpful but not practical in real-time.

The effects of censoring bias were more subtle on a daily scale. In our simulations, we found that failing to account for censoring typically led to biased estimates of the standard deviation but unbiased estimates of the mean. When inappropriate censoring adjustments were used the estimated mean was also biased. An example common in the literature of an inappropriate censoring adjustment is to use a single-day discretisation, which only accounts for censoring of a single event and induces a bias of half the unaccounted-for censoring interval to the mean of the estimated distribution. The approximate-latent-variable model, the interval-reduced-censoring-and-truncation model, the dynamical-bias-correction method, and the joint-modeling method gave relatively unbiased and precise estimates of underlying delay distribution parameters for the daily reporting scenario.

### 4.2 Limitations

While we have shown that accounting for both truncation and censoring biases is critical to accurately estimating epidemiological delay distributions, some methods can be more computationally costly than others. In particular, latent variable methods required nearly an order of magnitude more computational time in most cases compared to non-latent variable methods. The dynamical bias correction method and the joint model of primary incidence and the forward distribution had similar computational requirements to the latent model. However, in most instances, these requirements were manageable with typical research computing resources (i.e., laptop scale). In settings with time-varying and partially pooled delay distributions, this may no longer be the case and so non-latent approaches may be favoured despite the slight increase in bias.

In terms of accounting for dynamical biases, previous studies (Verity et al., 2020a; Britton and Scalia Tomba, 2019; Park et al., 2021) focused on the exponential growth phase, simplifying the problem. This approach is acceptable as long as growth stays roughly constant. However, propagating uncertainty appropriately is difficult with this approach and stable growth rates are rare in practice. Here, we present a novel version of the growth rate correction method that accounts for flexible changes in incidence patterns. Whilst this method performed well in retrospective settings, its application to real-time epidemics is currently limited due to its dependence on untruncated incidence data. It was routinely outperformed for real-time usage by other methods that directly accounted for truncation and censoring. These approaches are also more readily implementable using existing software, can account for censoring windows of varying sizes, and were more numerical stable.

Our case study of the Ebola virus disease epidemic revealed important gaps in the methods we present here. First, the reporting delays show considerable variation throughout the epidemic which is significantly larger than any bias due to censoring or truncation; current methods are not able to fully account for temporal changes in the delay distribution. While it may seem relatively straightforward to extend the model to allow for time-varying parameters across primary cohorts, censoring of the primary events complicates the problem by adding uncertainty to their cohort time. The dynamical correction method performed particularly poorly on the Ebola virus disease data. This was due to left truncation which was caused by dividing the data into four observation periods. Properly accounting for this induced left-truncation would require integrating the entire backward distribution, which is currently computationally impractical.

A key limitation of our work is that we only consider an idealized daily censoring process—in principle, our methodology and implementation support alternative censoring periods, except for the dynamic adjustment model. However, we did explore short delays and high growth rates which has a similar impact to having longer censoring intervals with long delays and slower growth rates. We found that exponential growth and truncation affect the distribution of event times within the censoring window, particularly when delay distributions are short relative to the length of the censoring window. These effects caused the empirical distribution of event times within the censoring interval to deviate from the assumed uniform distribution and we expect larger biases for wider censoring intervals. More work is needed to develop robust methods for dealing with wider censoring intervals that account for the underlying generative process of primary events. More generally the simulated scenarios we considered were idealised and did not include observation error for either the primary or secondary events. Testing our models against idealized scenarios allowed us to identify the detailed sources of biases, but may have favoured methods that did not try and account for observation error and other real-world sources of biases.

### 4.3 Generalisability

We only considered lognormal distributions in this paper for brevity and because it is commonly used in the literature, but our findings generalise across other distributions. Our implementations are also readily extensible to other distributions. We also did not consider mixtures of distributions, which can better describe some epidemiological delay distributions that are generated using multiple transmission or disease progression states (Vink et al., 2014). In addition, we do not include non-parametric or hazard-based methods in our assessment, although the joint incidence and forward distribution we did consider has been generalised to support these methods. However, again our key findings generalise to both these settings and, since our models are implemented using the `brms` package, it would be relatively easy to include these complexities with minimal additional work. Finally, the dynamical correction method assumes that the incidence is known exactly—a joint estimation of the incidence pattern and delay distribution, similar to that used in the joint incidence and forward distribution model, may improve this method’s real-time performance. Despite these limitations, our conclusions about the importance of truncation and censoring biases should be carefully considered for any epidemic analyses, especially when estimating delay distributions.

In this work, we have primarily focused on inferring distributions of non-transmission intervals (i.e., excluding generation- and serial-interval distributions). Although it would be possible to apply our methods to infer the mean and standard deviations of the transmission intervals, there are additional complications that we did not consider. In particular, transmission intervals may not be independent of each other if they share the same source case. Other problems include identifying intermediate hosts and the possibility of multiple potential source cases for an infectee. More work is needed to validate methods for inferring transmission interval distributions.

Estimation of epidemiological delay distributions is a common task in infectious disease modelling. In this work, we have given particular focus to daily censoring and right truncation adjustment as these are the most common scenarios researchers face when estimating delay distributions. When censoring is adjusted for, it is commonly assumed to be only censoring for the primary event and not the secondary event (i.e in the daily setting only account for a day vs two days of censoring). For example, researchers often account for the censoring in the infection time when estimating incubation-period distributions, but not in the symptom onset time. Right truncation is rarely adjusted for, and when it is, methods with limited theoretical support are commonly used which do not, or only partially, account for this bias. These methods are rarely validated against simulations (Backer et al., 2020; Linton et al., 2020) but are nonetheless often reused. For example, there has been an increased usage of methods that account for dynamical and right-truncation biases at the same time (Guo et al., 2023b,a); however, these two biases each pertain to backward and forward distributions, respectively and therefore should not be combined. Approaches that combine both biases will overcompensate for missing observations and overestimate the mean. As early estimates of epidemiological distributions are rarely re-estimated after the initial phase of an epidemic, due to lack of resources and the difficulty in collecting data, these biased estimates may remain the canonical ones throughout an epidemic, and beyond, further biasing decision-making.

More work is needed to improve software support for estimating distributions. Our code base from this work is now part of the epinowcast community, a group of infectious disease researchers aiming to improve epidemic and surveillance tools, meaning that it will be further developed into a robust tool. New members and support towards this aim are warmly welcomed. Further work is also needed to understand optimal methods for modelling time-varying distributions and mixture distributions with latent components where both may suffer from right truncation.

### 4.4 Conclusions

This study shows that care is required when estimating epidemiological distributions. We provide theory, methods, and tools to enable practitioners to circumvent common pitfalls that we have described and compare these methods in a range of simulated and real-world scenarios. Future epidemic analyses should carefully consider the different biases outlined in this study and make sure to use methods that can account for them and that have been robustly validated.

## Data Availability

All code used in the present study are available on https://github.com/parksw3/epidist-paper

## Acknowledgements

We thank Michael DeWitt for helpful comments on the manuscript. SF was supported by Wellcome Trust (210758/Z/18/Z).

## Supplementary Figures

**Figure 8:**
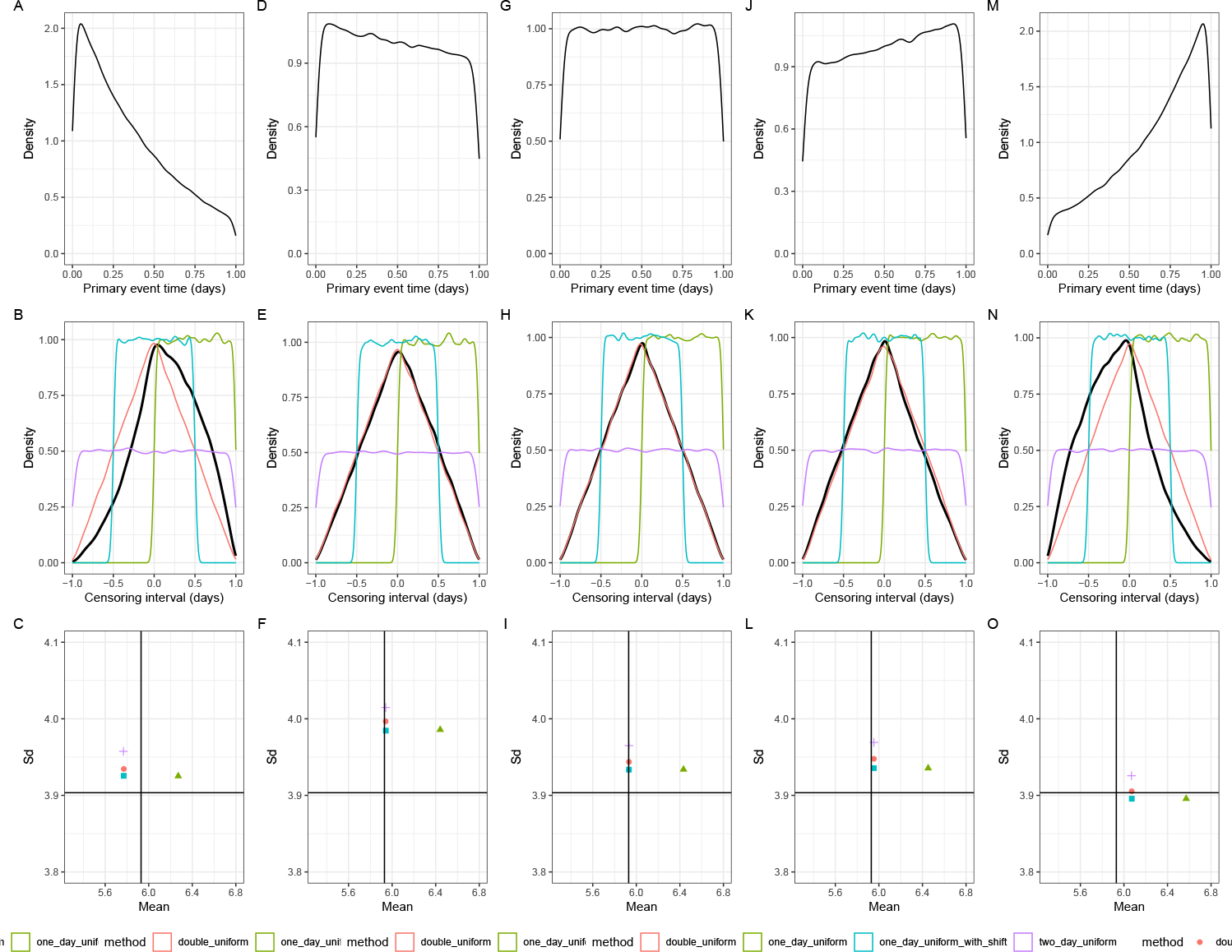
The impact of assumptions about prior distributions on converting discrete-time distributions to continuous-time distributions. (A, D, G, J, M) The distribution of primary event times within a one-day censoring interval across different growth rates. (B, E, H, K, N) The corresponding distribution of weights for the interval-reduced censoring (black lines) against different approximations (colored lines). (C, F, I, L, O) The means and standard deviations of the resulting continuous-time distributions across different assumptions (colored points) against the true mean (vertical lines) and standard deviations (horizontal lines).

**Figure 9:**
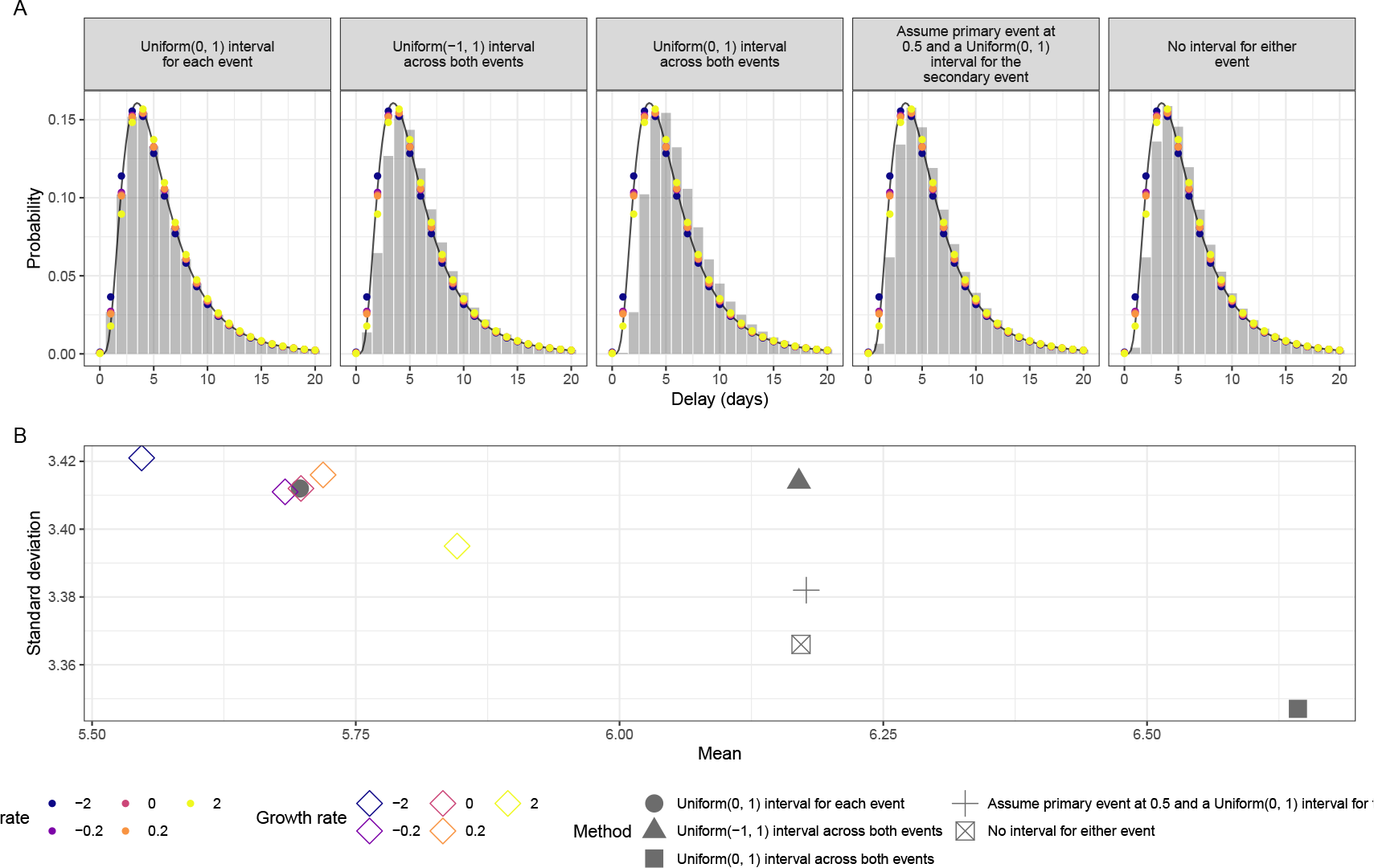
The impact of assumptions about prior distributions on converting continuous-time distributions to discrete-time probability mass functions. (A) Empirically observed probability mass functions (PMFs) from an underlying continuous lognormal distribution delay simulation (mean: 5.9 days, standard deviation: 3.9 days) with daily censoring. Observed PMFs for each interval-reduced censoring window approximation are shown using grey bars, PMFs under different growth rate assumptions for the primary interval are shown using coloured points. The black line indicates the underlying continuous probability density function used for simulation. (B) Empirically observed means and standard deviations from the same simulation as (D). Growth rate-adjusted primary censoring intervals are shown with coloured diamonds, and method-based censoring are indicated using grey shapes.

**Figure 10:**
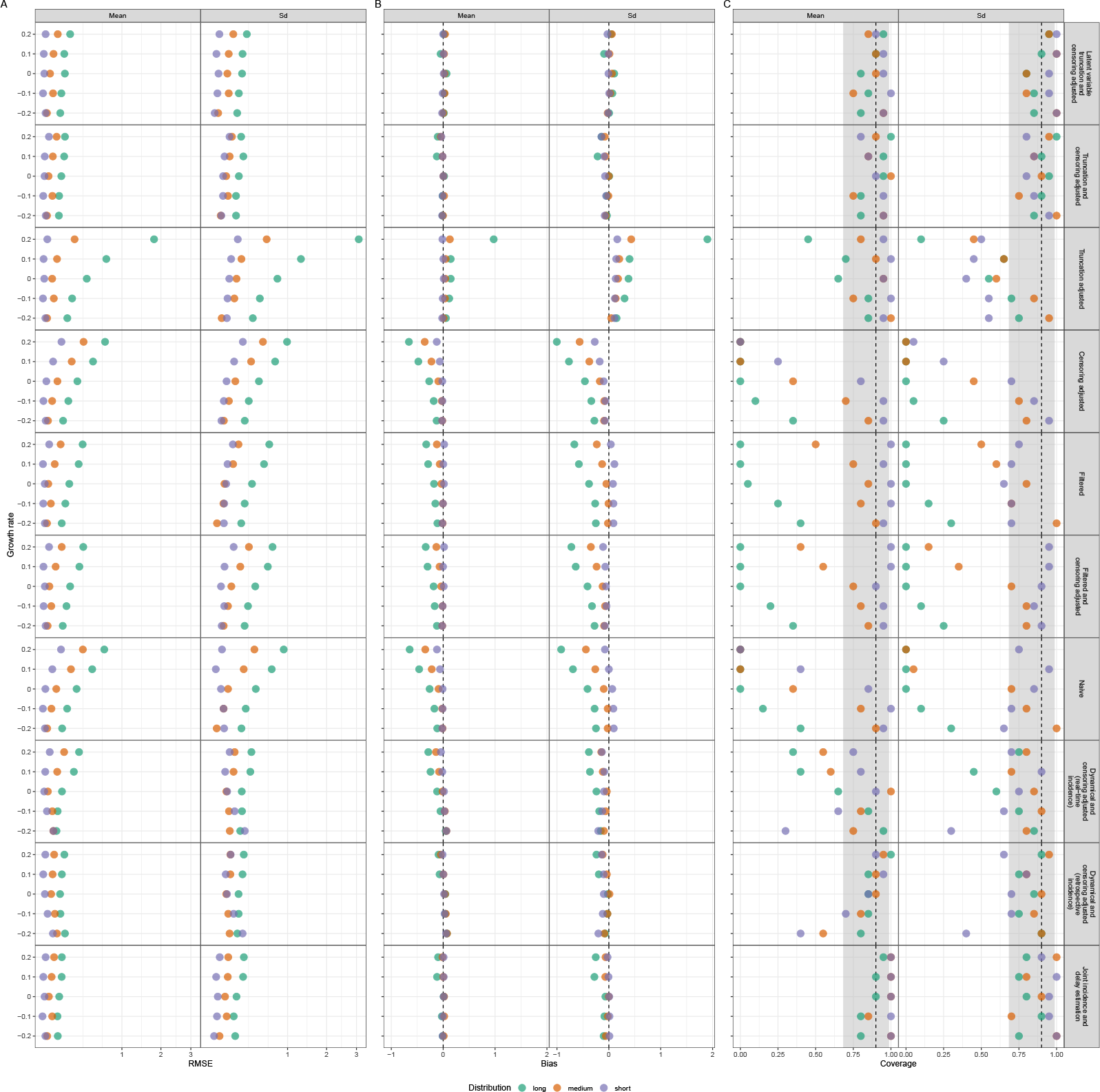
Coloured points represent summary statistics for each distribution scenario (short, medium, and long). (A) RMSE (root mean squared error) of each method. (B) Relative bias of each method. Vertical dashed lines represent the unbiased estimate. (C) Coverage probability of each method. Vertical dashed lines represent the 90% coverage probability. Shaded regions represent the 95% binomial confidence interval around 90% given number of simulations.

**Figure 11:**
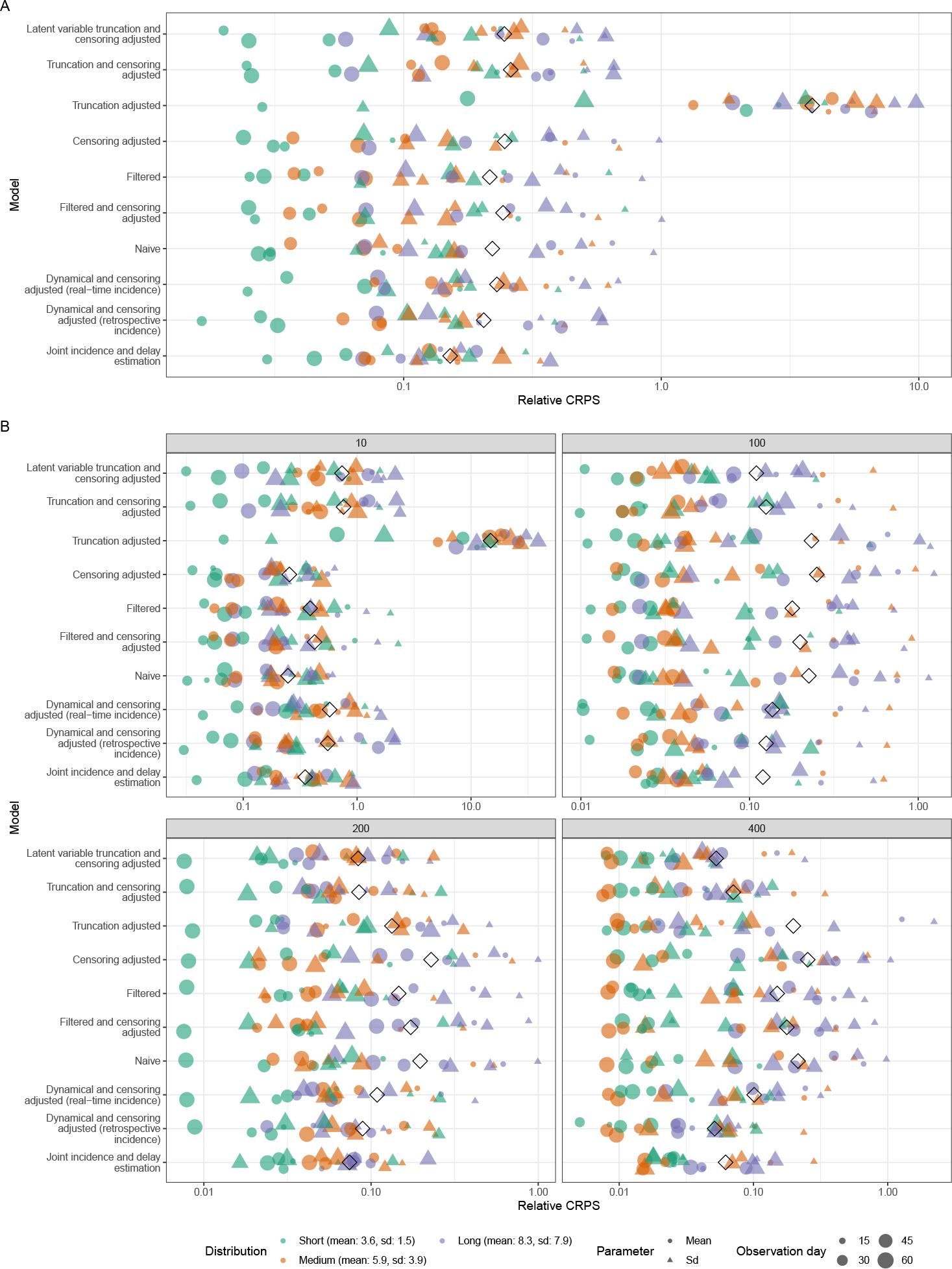
(A) Relative CRPS of each method across all sample sizes investigated (10, 100, 200, 400) for the outbreak scenario simulation. Here black diamonds represent the global relative CRPS. Coloured points represent the mean CRPS for each distribution scenario with the shape indicating if the score is for the mean (circle) or standard deviation (Sd, triangle). (B) Relative CRPS of each method stratified by sample sizes (10, 100, 200, 400) for the outbreak scenario simulation.

**Figure 12:**
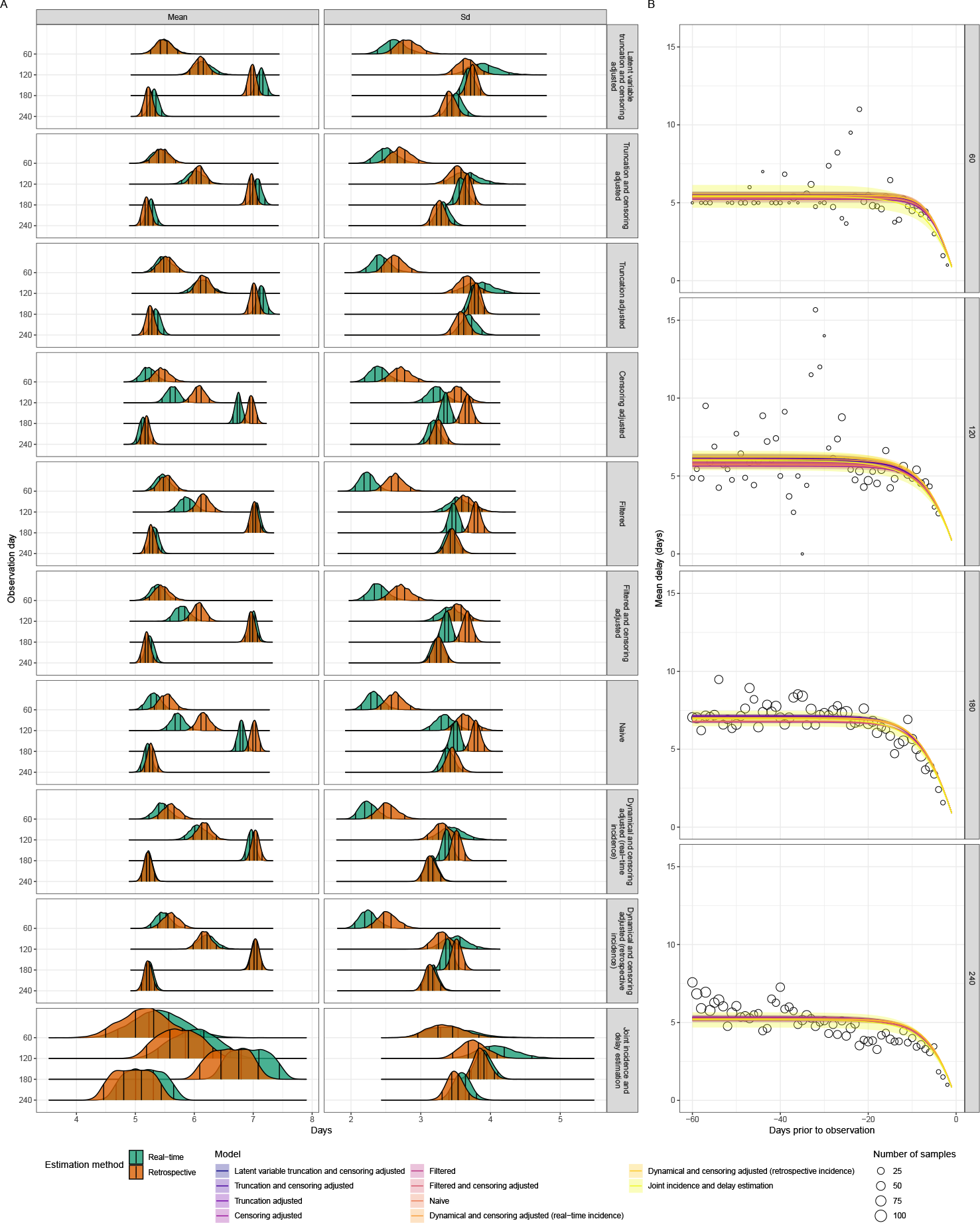
(A) Posterior distributions of mean and standard deviation (Sd). Vertical lines represent the 5%, 35%, 65%, and 95% quantiles respectively. Models are ordered based on the order used in the methods section. (B) Posterior predictions of truncated mean overtime against the observed forward mean. Lines and shaded regions represent posterior median and 95% credible intervals. Points represent the observed mean values. Note that some models are over-plotted here and hence may not be clearly distinguishable.

## References

Abbott, S., Hellewell, J., Thompson, R. N., Sherratt, K., Gibbs, H. P., Bosse, N. I., Munday, J. D., Meakin, S., Doughty, E. L., Chun, J. Y., Chan, Y.-W. D., Finger, F., Campbell, P., Endo, A., Pearson, C. A. B., Gimma, A., Russell, T., Flasche, S., Kucharski, A. J., Eggo, R. M., Funk, S., and CMMID COVID modelling group (2020). Estimating the time-varying reproduction number of SARS-CoV-2 using national and subnational case counts. Wellcome Open Res., 5:112.

Abbott, S., Lison, A., and Funk, S. (2021). epinowcast: Flexible hierarchical now-casting. Zenodo.

Backer, J. A., Eggink, D., Andeweg, S. P., Veldhuijzen, I. K., van Maarseveen, N., Vermaas, K., Vlaemynck, B., Schepers, R., van den Hof, S., Reusken, C. B., et al. (2022). Shorter serial intervals in SARS-CoV-2 cases with Omicron BA. 1 variant compared with Delta variant, the Netherlands, 13 to 26 December 2021. Eurosurveillance, 27(6):2200042.

Backer, J. A., Klinkenberg, D., and Wallinga, J. (2020). Incubation period of 2019 novel coronavirus (2019-nCoV) infections among travellers from Wuhan, China, 20–28 January 2020. Eurosurveillance, 25(5):2000062.

Beesley, L. J., Osthus, D., and Del Valle, S. Y. (2022). Addressing delayed case reporting in infectious disease forecast modeling. PLoS Comput. Biol., 18(6):e1010115.

Betancourt, M. (2017). Diagnosing biased inference with divergences. Stan Case Studies, 4.

Boettiger, C. (2015). An introduction to Docker for reproducible research. ACM SIGOPS Operating Systems Review, 49(1):71–79.

Bosse, N. I., Abbott, S., Cori, A., van Leeuwen, E., Bracher, J., and Funk, S. (2023). Transformation of forecasts for evaluating predictive performance in an epidemiological context.

Bosse, N. I., Gruson, H., Cori, A., van Leeuwen, E., Funk, S., and Abbott, S. (2022). Evaluating forecasts with scoringutils in r. arXiv.

Britton, T. and Scalia Tomba, G. (2019). Estimation in emerging epidemics: biases and remedies. Journal of the Royal Society Interface, 16(150):20180670.

Brookmeyer, R. and Damiano, A. (1989). Statistical methods for short-term projections of AIDS incidence. Statistics in Medicine, 8(1):23–34.

Bürkner, P.-C. (2018). Advanced Bayesian multilevel modeling with the R package brms. The R Journal, 10(1):395–411.

Cain, K. C., Harlow, S. D., Little, R. J., Nan, B., Yosef, M., Taffe, J. R., and Elliott, M. R. (2011). Bias due to left truncation and left censoring in longitudinal studies of developmental and disease processes. American journal of epidemiology, 173(9):1078–1084.

Champredon, D. and Dushoff, J. (2015). Intrinsic and realized generation intervals in infectious-disease transmission. Proceedings of the Royal Society B: Biological Sciences, 282(1821):20152026.

Cori, A., Ferguson, N. M., Fraser, C., and Cauchemez, S. (2013). A new framework and software to estimate time-varying reproduction numbers during epidemics. American journal of epidemiology, 178(9):1505–1512.

Fang, L.-Q., Yang, Y., Jiang, J.-F., Yao, H.-W., Kargbo, D., Li, X.-L., Jiang, B.-G., Kargbo, B., Tong, Y.-G., Wang, Y.-W., Liu, K., Kamara, A., Dafae, F., Kanu, A., Jiang, R.-R., Sun, Y., Sun, R.-X., Chen, W.-J., Ma, M.-J., Dean, N. E., Thomas, H., Longini, Jr, I. M., Halloran, M. E., and Cao, W.-C. (2016). Transmission dynamics of ebola virus disease and intervention effectiveness in sierra leone. Proc. Natl. Acad. Sci. U. S. A., 113(16):4488–4493.

Flaxman, S., Mishra, S., Gandy, A., Unwin, H. J. T., Mellan, T. A., Coupland, H., Whittaker, C., Zhu, H., Berah, T., Eaton, J. W., Monod, M., Imperial College COVID-19 Response Team, Ghani, A. C., Donnelly, C. A., Riley, S., Vollmer, M. A. C., Ferguson, N. M., Okell, L. C., and Bhatt, S. (2020). Estimating the effects of non-pharmaceutical interventions on COVID-19 in europe. Nature, 584(7820):257– 261.

Fraser, C., Donnelly, C. A., Cauchemez, S., Hanage, W. P., Van Kerkhove, M. D., Hollingsworth, T. D., Griffin, J., Baggaley, R. F., Jenkins, H. E., Lyons, E. J., et al. (2009). Pandemic potential of a strain of influenza A (H1N1): early findings. science, 324(5934):1557–1561.

Fraser, C., Riley, S., Anderson, R. M., and Ferguson, N. M. (2004). Factors that make an infectious disease outbreak controllable. Proceedings of the National Academy of Sciences, 101(16):6146–6151.

Gabry, J. and Češnovar, R. (2021). cmdstanr: R Interface to ‘CmdStan’. https://mc-stan.org/cmdstanr,

https://discourse.mc-stan.org.

Gelman, A., Carlin, J. B., Stern, H. S., Dunson, D. B., Vehtari, A., and Rubin, D. B. (2013). Bayesian data analysis. CRC press.

Gelman, A. and Rubin, D. B. (1992). Inference from iterative simulation using multiple sequences. Statistical science, 7(4):457–472.

Ghani, A. C., Donnelly, C. A., Cox, D. R., Griffin, J. T., Fraser, C., Lam, T. H., Ho, L. M., Chan, W. S., Anderson, R. M., Hedley, A. J., and Leung, G. M. (2005). Methods for estimating the case fatality ratio for a novel, emerging infectious disease. Am. J. Epidemiol., 162(5):479–486.

Gneiting, T. and Raftery, A. E. (2007). Strictly Proper Scoring Rules, Prediction, and Estimation. Journal of the American Statistical Association, 102(477):359– 378. DOI: 10.1198/016214506000001437.

Gostic, K., Gomez, A. C., Mummah, R. O., Kucharski, A. J., and Lloyd-Smith, J. O. (2020). Estimated effectiveness of symptom and risk screening to prevent the spread of COVID-19. Elife, 9.

Günther, F., Bender, A., Katz, K., Küchenhoff, H., and Höhle, M. (2021). Nowcast-ing the COVID-19 pandemic in Bavaria. 63(3):490–502.

Guo, Z., Zhao, S., Sun, S., He, D., Chong, K. C., and Yeoh, E. K. (2023a). Estimation of the serial interval of monkeypox during the early outbreak in 2022. Journal of Medical Virology, 95(1):e28248.

Guo, Z., Zhao, S., Yam, C. H. K., Li, C., Jiang, X., Chow, T. Y., Chong, K. C., and Yeoh, E. K. (2023b). Estimating the serial intervals of SARS-CoV-2 Omicron BA. 4, BA. 5, and BA. 2.12. 1 variants in Hong Kong. Influenza and Other Respiratory Viruses, 17(2):e13105.

Hart, W. S., Maini, P. K., and Thompson, R. N. (2021). High infectiousness immediately before COVID-19 symptom onset highlights the importance of continued contact tracing. Elife, 10:e65534.

He, X., Lau, E. H., Wu, P., Deng, X., Wang, J., Hao, X., Lau, Y. C., Wong, J. Y., Guan, Y., Tan, X., et al. (2020). Temporal dynamics in viral shedding and transmissibility of COVID-19. Nature medicine, 26(5):672–675.

Hellewell, J., Abbott, S., Gimma, A., Bosse, N. I., Jarvis, C. I., Russell, T. W., Munday, J. D., Kucharski, A. J., Edmunds, W. J., Sun, F., et al. (2020). Feasibility of controlling COVID-19 outbreaks by isolation of cases and contacts. The Lancet Global Health, 8(4):e488–e496.

Höhle, M. and an der Heiden, M. (2014). Bayesian nowcasting during the STEC O104:h4 outbreak in Germany, 2011. 70(4):993–1002.

Kalbfleisch, J. D. and Lawless, J. F. (1989). Inference based on retrospective ascertainment: an analysis of the data on transfusion-related AIDS. Journal of the American Statistical Association, 84(406):360–372.

Landau, W. M. (2021). The targets r package: a dynamic make-like function-oriented pipeline toolkit for reproducibility and high-performance computing. Journal of Open Source Software, 6(57):2959.

Lauer, S. A., Grantz, K. H., Bi, Q., Jones, F. K., Zheng, Q., Meredith, H. R., Azman, A. S., Reich, N. G., and Lessler, J. (2020). The incubation period of coronavirus disease 2019 (COVID-19) from publicly reported confirmed cases: estimation and application. Annals of internal medicine, 172(9):577–582.

Lindsey, J. C. and Ryan, L. M. (1998). Methods for interval-censored data. Statistics in medicine, 17(2):219–238.

Linton, N. M., Kobayashi, T., Yang, Y., Hayashi, K., Akhmetzhanov, A. R., Jung, S.-m., Yuan, B., Kinoshita, R., and Nishiura, H. (2020). Incubation period and other epidemiological characteristics of 2019 novel coronavirus infections with right truncation: a statistical analysis of publicly available case data. Journal of clinical medicine, 9(2):538.

Lipsitch, M., Donnelly, C. A., Fraser, C., Blake, I. M., Cori, A., Dorigatti, I., Ferguson, N. M., Garske, T., Mills, H. L., Riley, S., Van Kerkhove, M. D., and Hernán, M. A. (2015). Potential biases in estimating absolute and relative Case-Fatality risks during outbreaks. PLoS Negl. Trop. Dis., 9(7):e0003846.

Lipsitch, M., Joshi, K., and Cobey, S. E. (2020). Comment on Pan A, Liu L, Wang C, et al: Association of Public Health Interventions With the Epidemiology of the COVID-19 Outbreak in Wuhan, China. JAMA.

Lison, A., Abbott, S., Huisman, J., and Stadler, T. (2023). Generative Bayesian modeling to nowcast the effective reproduction number from line list data with missing symptom onset dates. arXiv.

Madewell, Z. J., Charniga, K., Masters, N. B., Asher, J., Fahrenwald, L., Still, W., Chen, J., Kipperman, N., Bui, D., Shea, M., Saunders, K., Saathoff-Huber, L., Johnson, S., Harbi, K., Berns, A. L., Perez, T., Gateley, E., Spicknall, I. H., Nakazawa, Y., Gift, T. L., and 2022 Mpox Outbreak Response Team (2023). Serial interval and incubation period estimates of monkeypox virus infection in 12 jurisdictions, united states, May-August 2022. Emerg. Infect. Dis., 29(4):818–821.

Marinović, A. B., Swaan, C., van Steenbergen, J., and Kretzschmar, M. (2015). Quantifying reporting timeliness to improve outbreak control. Emerging infectious diseases, 21(2):209.

Miura, F., van Ewijk, C. E., Backer, J. A., Xiridou, M., Franz, E., de Coul, E. O., Brandwagt, D., van Cleef, B., van Rijckevorsel, G., Swaan, C., et al. (2022). Estimated incubation period for monkeypox cases confirmed in the Netherlands, May 2022. Eurosurveillance, 27(24):2200448.

Nolen, L. D., Osadebe, L., Katomba, J., Likofata, J., Mukadi, D., Monroe, B., Doty, J., Hughes, C. M., Kabamba, J., Malekani, J., et al. (2016). Extended human-to-human transmission during a monkeypox outbreak in the Democratic Republic of the Congo. Emerging infectious diseases, 22(6):1014.

Overton, C. E., Pellis, L., Stage, H. B., Scarabel, F., Burton, J., Fraser, C., Hall, I., House, T. A., Jewell, C., Nurtay, A., et al. (2022). EpiBeds: Data informed modelling of the COVID-19 hospital burden in England. PLoS Computational Biology, 18(9):e1010406.

Pan, C., Cai, B., and Wang, L. (2020). A bayesian approach for analyzing partly interval-censored data under the proportional hazards model. Statistical methods in medical research, 29(11):3192–3204.

Park, S. W., Sun, K., Abbott, S., Sender, R., Bar-On, Y. M., Weitz, J. S., Funk, S., Grenfell, B., Backer, J. A., Wallinga, J., et al. (2022). Inferring the differences in incubation-period and generation-interval distributions of the Delta and Omicron variants of SARS-CoV-2. medRxiv, pages 2022–07.

Park, S. W., Sun, K., Champredon, D., Li, M., Bolker, B. M., Earn, D. J., Weitz, J. S., Grenfell, B. T., and Dushoff, J. (2021). Forward-looking serial intervals correctly link epidemic growth to reproduction numbers. Proceedings of the National Academy of Sciences, 118(2):e2011548118.

R Core Team (2019). R: A Language and Environment for Statistical Computing. R Foundation for Statistical Computing, Vienna, Austria.

Reich, N. G., Lessler, J., and Azman, A. S. (2010). coarseDataTools: A collection of functions to help with analysis of coarsely observed data. R package version 0. 6–6.

Reich, N. G., Lessler, J., Cummings, D. A., and Brookmeyer, R. (2009). Estimating incubation period distributions with coarse data. Statistics in medicine, 28(22):2769–2784.

Seaman, S. R., Presanis, A., and Jackson, C. (2022). Estimating a time-to-event distribution from right-truncated data in an epidemic: a review of methods. Statistical methods in medical research, 31(9):1641–1655.

Sender, R., Bar-On, Y., Park, S. W., Noor, E., Dushoff, J., and Milo, R. (2022). The unmitigated profile of COVID-19 infectiousness. Elife, 11:e79134.

Singanayagam, A., Patel, M., Charlett, A., Bernal, J. L., Saliba, V., Ellis, J., Ladhani, S., Zambon, M., and Gopal, R. (2020). Duration of infectiousness and correlation with RT-PCR cycle threshold values in cases of COVID-19, England, January to May 2020. Eurosurveillance, 25(32):2001483.

Stan Development Team (2020). Prior Choice Recommendations. https://github.com/stan-dev/stan/wiki/Prior-Choice-Recommendations.

Stan Development Team (2021). Stan Modeling Language Users Guide and Reference Manual, 2.28.1. https://mc-stan.org.

Sun, J. (1995). Empirical estimation of a distribution function with truncated and doubly interval-censored data and its application to AIDS studies. Biometrics, pages 1096–1104.

Svensson, A. (2007). A note on generation times in epidemic models. Math. Biosci., 208(1):300–311.

Thompson, R. N., Stockwin, J. E., van Gaalen, R. D., Polonsky, J. A., Kamvar, Z. N., Demarsh, P. A., Dahlqwist, E., Li, S., Miguel, E., Jombart, T., Lessler, J., Cauchemez, S., and Cori, A. (2019). Improved inference of time-varying reproduction numbers during infectious disease outbreaks. Epidemics, 29:100356.

Tindale, L. C., Stockdale, J. E., Coombe, M., Garlock, E. S., Lau, W. Y. V., Saraswat, M., Zhang, L., Chen, D., Wallinga, J., and Colijn, C. (2020). Evidence for transmission of COVID-19 prior to symptom onset. Elife, 9:e57149.

Ushey, K. (2021). renv: Project Environments. R package version 0.14.0.

Verity, R., Okell, L. C., Dorigatti, I., Winskill, P., Whittaker, C., Imai, N., Cuomo-Dannenburg, G., Thompson, H., Walker, P. G., Fu, H., et al. (2020a). Estimates of the severity of coronavirus disease 2019: a model-based analysis. The Lancet infectious diseases, 20(6):669–677.

Verity, R., Okell, L. C., Dorigatti, I., Winskill, P., Whittaker, C., Imai, N., Cuomo-Dannenburg, G., Thompson, H., Walker, P. G., Fu, H., et al. (2020b). Estimates of the severity of coronavirus disease 2019: a model-based analysis. The Lancet infectious diseases, 20(6):669–677.

Verity, R., Okell, L. C., Dorigatti, I., Winskill, P., Whittaker, C., Imai, N., Cuomo-Dannenburg, G., Thompson, H., Walker, P. G. T., Fu, H., Dighe, A., Griffin, J. T., Baguelin, M., Bhatia, S., Boonyasiri, A., Cori, A., Cucunubá, Z., FitzJohn, R., Gaythorpe, K., Green, W., Hamlet, A., Hinsley, W., Laydon, D., Nedjati-Gilani, G., Riley, S., van Elsland, S., Volz, E., Wang, H., Wang, Y., Xi, X., Donnelly, C. A., Ghani, A. C., and Ferguson, N. M. (2020c). Estimates of the severity of coronavirus disease 2019: a model-based analysis. Lancet Infect. Dis., 20(6):669–677.

Vink, M. A., Bootsma, M. C. J., and Wallinga, J. (2014). Serial intervals of respiratory infectious diseases: a systematic review and analysis. American journal of epidemiology, 180(9):865–875.

Ward, T., Christie, R., Paton, R. S., Cumming, F., and Overton, C. E. (2022). Transmission dynamics of monkeypox in the United Kingdom: contact tracing study. bmj, 379.

Ward, T. and Johnsen, A. (2021). Understanding an evolving pandemic: An analysis of the clinical time delay distributions of COVID-19 in the United Kingdom. PLoS One, 16(10):e0257978.

Xin, H., Wong, J. Y., Murphy, C., Yeung, A., Taslim Ali, S., Wu, P., and Cowling, B. J. (2021). The incubation period distribution of coronavirus disease 2019: a systematic review and meta-analysis. Clinical Infectious Diseases, 73(12):2344–2352.

